# Same-day hybrid capture workflow for diagnosis and surveillance of respiratory pathogens

**DOI:** 10.64898/2026.10.09.26365131

**Authors:** Laura Buggiotti, Amelie Busam, Luz Marina Martin Bernal, Tony Brooks, Sarah Buddle, Angelika Kopec, Lamisha Ahmed, Gabriela Kuczmar, Alexander Lennon, Sergi Castellano, Peter Simmonds, Heli Harvala, Tanya Golubchik, Julianne R Brown, Rachel Williams, Judith Breuer, Sofia Morfopoulou, Oscar Enrique Torres Montaguth

## Abstract

Hybrid capture approaches can detect a broad range of pathogens directly from clinical samples with greater sensitivity than untargeted approaches. Clinical adoption has been limited by long turnaround times, complex workflows, and perceived high costs. We iteratively developed and validated a rapid hybrid capture workflow based on Illumina’s RPIP protocol (standard workflow). Rapid-RPIP v1 reduced wet-lab time from ∼11.4 to ∼7.3 h, for a turnaround time (TAT) of <15 h. Rapid-RPIP v2 further reduced wet-lab time by ∼0.7 h and used Illumina Turbo Sequencing shortening the sequencing time from ∼5.5 to ∼2.5 h thus achieving a TAT of <10 h without compromising sensitivity and yielding higher RPKMs for RNA viruses than the standard workflow. This approach could be readily adapted to a broad range of clinical and public health applications.

## Background

Respiratory tract infections (RTIs) remain a major cause of global morbidity and mortality, particularly among vulnerable populations such as children and immunocompromised individuals [1]. Given the wide diversity of pathogens associated with RTIs, approximately 15–30% of cases remain undiagnosed, which may limit effective treatment and infection control [2,3]. Conventional diagnostic methods, such as culture and qPCR panels, detect only pathogens that are culturable or that have been pre-specified respectively, restricting their ability to identify novel or unexpected organisms. Furthermore, sequential testing using different assays delays diagnosis and appropriate clinical intervention [4].

Clinical metagenomics enables sequencing of all genetic material within a sample, allowing agnostic pathogen detection in a single assay [5,6]. It can also provide valuable information on antimicrobial resistance (AMR) and virulence factors. Metagenomics has been mainly applied in clinical settings for the diagnosis of encephalitis and meningitis, fever of unknown origin and sepsis [7–10]. However, its widespread implementation has been limited by reduced sensitivity compared to established molecular diagnostic methods, long turnaround times, high costs, workflow complexity and challenges in the interpretation of results arising from the presence of commensal organisms or background contamination [5,6].

Recently, rapid metagenomic workflows for detecting respiratory infections have been developed and validated [9–12,34]. These approaches can deliver results in a single day, with preliminary findings available within approximately 7–8 h and final reports within 24 h. These workflows use differential lysis to reduce host material, improving sensitivity in samples with a high host background. However, this approach can bias detection toward DNA pathogens and may negatively affect the detection of RNA viruses and intracellular pathogens [6,13,14]. In addition, depletion of host material limits the possibility of using host genomic and transcriptional biomarkers, which may provide additional information for clinical interpretation and patient management [15–17].

Hybrid capture workflows enable targeted enrichment and detection of hundreds of pathogens directly from clinical samples in a single test. These methods use probes complementary to target genomic sequences to selectively enrich those sequences prior to sequencing, offering 10- to 100-fold higher sensitivity than untargeted approaches. [18,19]. Commercially available workflows, such as the Illumina Respiratory Pathogen ID/AMR Enrichment Panel (RPIP), have been designed for respiratory infections with the RPIP panel targeting over 280 pathogens, including viruses, bacteria, and fungi, as well as associated antimicrobial resistance markers [20]. However, the complexity and duration of these protocols, which require more than 12 h for the wet-lab component, have limited their implementation in clinical settings [21].

To overcome this limitation, we have developed and validated a rapid hybrid capture protocol using the RPIP. Through iterative optimisation using both commercial respiratory mock community controls and residual clinical samples, we have refined the workflow from nucleic acid extraction to sequencing and evaluated its performance on respiratory samples in comparison with standard-of-care (SOC) testing. The optimised workflow enables sample-to-result turnaround in under 10 h, delivering same-day comprehensive pathogen detection with performance comparable to routine diagnostic methods. This establishes a core workflow that can be adapted to other sample types and infectious disease syndromes.

## Methods

### Mock Community Controls

The NATtrol Respiratory Panel 2.1 (RP2.1; ZeptoMetrix, NATRPC2.1-BIO) was used as mock communities to assess the detection capabilities of both standard and rapid sequencing workflows. RP2.1 contains two defined mixtures (Control 1 and Control 2) of purified bacterial cells and viral particles representing common respiratory pathogens (Table 1). The material is chemically modified to eliminate infectivity. Each control was processed neat and at 10-fold dilution through both workflows.

**Table 1.**
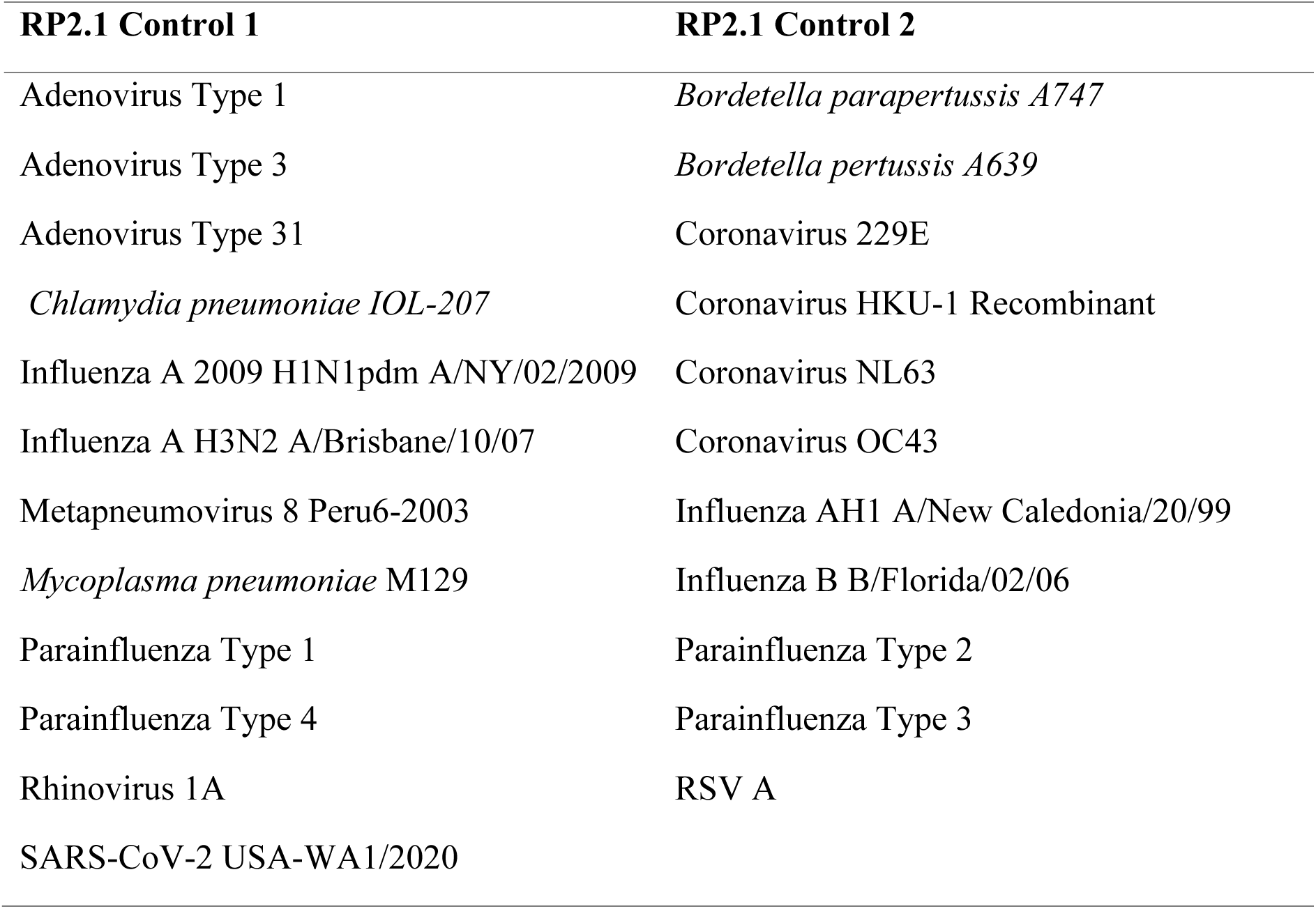
Composition of NATtrol RP2.1 mock community controls.

### Clinical Samples

Residual clinical respiratory specimens (nasopharyngeal aspirates and endotracheal secretions) from patients with suspected respiratory tract infection were processed in parallel through both the Standard RPIP and the newly developed Rapid-RPIP sequencing workflows (Table S1). Samples were tested for bacteria and viruses using SOC diagnostic methods. Where available, aliquots from the same sample that underwent diagnostic bacterial culture and PCR were used for metagenomic sequencing. Respiratory cultures were performed using MacConkey agar, Columbia Blood, Chocolate Blood and Sabouraud agar. PCR was performed using the QIAstat-DX Respiratory panel or an in-house PCR panel accredited to ISO:15189 (2022) standards (that targets Influenza A+B, SARS-CoV-2, Parainfluenza type 1, parainfluenza 1+2+3, RSV A/B, Human Metapneumovirus, Rhinovirus A and Adenovirus). If the sample volume available was insufficient to perform all tests, results from samples collected within ±2 days of the metagenomic sample collection date were included. Specific PCRs were performed to confirm additional pathogens identified by RPIP.

### Respiratory sample storage and initial lysis step

A total of 500 μL of sample was mixed with extraction reaction mix (498 μL 2× DNA/RNA Shield (Zymo Research, R1200) and 2μL MS2 internal positive control) and stored at −80°C until processing. Samples were thawed on ice, and 800 μL of the resulting mixture was transferred to a ZR BashingBead Lysis Tube (0.1- and 0.5-mm beads; Zymo Research, S6012-50) and lysed using a FastPrep-24™ 5G bead beater with two cycles of 30 s at 6 m/s separated by a 5 min interval. Lysates were then processed using the RPIP workflows described below. Detailed information, including preparation of the MS2 internal positive control, is provided in Protocols S1 and S2.

### Standard RPIP Workflow

Samples were processed using the Respiratory Pathogen Identification/Antimicrobial Resistance Panel (RPIP; Illumina, 20047050) according to the manufacturer’s instructions. Briefly, total nucleic acid extraction was performed using the ZymoBIOMICS DNA/RNA Miniprep Kit (Zymo Research, R2002). Following nucleic acid purification, RNA underwent DNase treatment before elution and cDNA synthesis. Extracted DNA and cDNA were subsequently combined and used for library preparation.

Sequencing libraries were prepared using the Illumina RNA Prep with Enrichment workflow, including tagmentation, PCR amplification, and indexing. Libraries were then pooled (three-plex) and enriched using the RPIP respiratory pathogen and antimicrobial resistance probe panel. Probe hybridisation was performed for 90 min at 58°C, followed by streptavidin bead capture, post-enrichment PCR amplification, and final library purification. Enriched libraries were quantified, normalised, pooled, and sequenced using the MiniSeq Rapid Reagent Kit (100 cycles; Illumina, 20044338).

### Rapid-RPIP v1 Workflow

The Rapid-RPIP v1 workflow was developed by adapting the Standard RPIP workflow, incorporating alternative methods and modifying the protocol with the aim of reducing workflow time without compromising performance. To this end, the ZymoBIOMICS DNA/RNA Miniprep Kit for nucleic acid extraction was replaced with the Quick-DNA/RNA Pathogen Miniprep Kit (Zymo Research, R1042), and samples were processed according to the manufacturer’s instructions.

To evaluate the performance of both extraction kits, we used the ZymoBIOMICS Microbial Community Standard (Zymo Research, D6300). Library preparation of the extracts was performed using the NEBNext Ultra II FS DNA Library Prep Kit (New England Biolabs, E7805) and sequenced using MiSeq Reagent Kit v2 (300 cycles; Illumina, MS-102-2002)

For cDNA synthesis, the RPIP cDNA synthesis workflow was replaced with a rapid protocol based on LunaScript RT SuperMix (New England Biolabs, E3010) for first-strand synthesis and Sequenase Version 2.0 DNA Polymerase (Thermo Fisher Scientific, 70775Y) for second-strand synthesis. Following cDNA synthesis, the standard bead-based clean-up step was replaced with enzyme heat inactivation. Detailed reaction conditions are provided in the Supplementary Methods.

Library preparation continued using the Illumina RNA Prep with Enrichment workflow; however, the extension step during the 17-cycle pre-hybridisation PCR was reduced from 60 s to 15 s at 72°C. During target enrichment, the hybridisation time at 58°C was reduced from 90 min to 30 min. All subsequent enrichment, amplification, and clean-up steps were performed according to the manufacturer’s instructions. Enriched libraries were quantified, normalised, pooled, and sequenced using the MiniSeq Rapid Reagent Kit (100 cycles; Illumina, 20044338). A complete step-by-step Rapid-RPIP v1 protocol is provided as Protocol S1.

### Rapid-RPIP v2 Workflow

The Rapid-RPIP v2 workflow was based on the Rapid-RPIP v1 workflow. Following the release of the Illumina Rapid Respiratory Pathogen Identification/Antimicrobial Resistance Panel (Illumina, 20158828), which incorporates an integrated cDNA synthesis and tagmentation approach, this workflow was adopted in place of the standalone cDNA synthesis protocol used in Rapid-RPIP v1. All other modifications introduced into the Rapid-RPIP v1 workflow, including the use of the Quick-DNA/RNA Pathogen Miniprep Kit for nucleic acid extraction, reduced PCR extension times, and a 30 min hybridisation step, were retained.

Enriched libraries were quantified, normalised, pooled, and sequenced using the MiSeq i100 Series 25M Reagent Kit (100 cycles; Illumina, 20126567) with Turbo Sequencing enabled. A complete step-by-step Rapid-RPIP V2 protocol is provided as Protocol S2.

### Bioinformatic Analysis

#### DRAGEN Microbial Enrichment Plus

All samples were analysed with DRAGEN Microbial Enrichment Plus v1.1.1 (DME+; Illumina Inc., San Diego, CA) on the BaseSpace Sequence Hub platform. Default pipeline settings were used throughout, including Illumina’s proprietary quality control thresholds, read filtering parameters and curated microbial reference databases. To assess organisms detected below the default reporting threshold, the secondary analysis option “Report microorganisms and/or AMR markers that are below threshold” was enabled. DME+ assigns each positive target to one of three phenotypic categories. Group 1 corresponds to microorganisms that are frequently considered part of the normal flora, colonizers, or contaminants but which may be associated with disease in certain settings. Group 2 corresponds to microorganisms that may represent normal flora, colonizers, or contaminants but are frequently associated with disease. Finally, group 3 corresponds to microorganisms that are not generally considered part of the normal flora, colonizers, or contaminants and are generally considered to be associated with disease.

#### Independent taxonomic classification

To provide context beyond the proprietary DME+ workflow, all samples additionally underwent taxonomic classification with nf-core/taxprofiler v1.2.4 [22], executed with Nextflow v25.04.6[23]. The pipeline included quality control with FastQC [24], adapter trimming with fastp [25], host read removal by Bowtie2[26] alignment to the human reference genome (GRCh38, Ensembl release 107), taxonomic classification with Kraken2 [27], Bayesian abundance re-estimation with Bracken [28], and output standardisation with taxpasta [29].

### Sensitivity

Sensitivity was assessed against routine SOC testing, namely culture and multiplex PCR as the reference standard, applying the specimen-type and collection-window criteria described above. Sensitivity was calculated as the proportion of SOC-positive organisms detected by each protocol (True Positive/ (True Positive +False Negative)), with Wilson 95% confidence intervals. Additional organisms detected that were not part of the routine SOC-positive reference set were tested by specific PCRs. Where the PCR was positive, the pathogen was classified as independently confirmed; PCR negatives were classified as unconfirmed and considered as false positives.

### Statistical Analysis

Concordance between protocols was assessed using Spearman rank correlation on raw RPKM (Reads per Kilobase Million) values and Pearson correlation on log₁₀-transformed RPKM. Pairwise log₂ fold changes (Rapid-RPIP v1/Standard or Rapid-RPIP v2/Rapid-RPIP v1) were computed per pathogen and compared between RNA and DNA pathogens using the Mann-Whitney U test. Differences in sensitivity between protocols were assessed by Fisher’s exact test. All analyses were conducted in R v4.3. A two-sided p-value < 0.05 was considered statistically significant; given the exploratory nature of the comparisons, no formal multiple-testing correction was applied.

### Validation of Kraken2/Bracken discordant calls

Pathogen sample pairs where Kraken2/Bracken called an organism positive and DME+ called it negative were identified and subjected to two-step validation. First, host-depleted reads were mapped to the correspondent pathogen reference genomes using BWA v0.7.17 (e.g. *Candida albicans* SC5314, GCF_000182965.3; CMV Merlin strain, NC_006273.2; Enterovirus CoxA16/CoxA8, U05876/AY421766*; Pseudomonas aeruginosa* PAO1, NC_002516.2; *Staphylococcus aureus* MRSA252, NC_002745.2), and coverage breadth and mean depth were calculated using samtools v1.14 in 500 bp windows. Second, reads classified by Kraken2 to each discordant organism were extracted using their taxonomy IDs, and the top 10 reads per organism were queried against the NCBI nr/nt database using BLASTn. Organisms were classified as genuine detections if BLASTn top hits showed ≥95% query coverage and ≥95% identity to the expected species; calls were classified as false positives where query coverage was substantially reduced (<95%) or top hits matched multiple unrelated taxa, indicating reads from conserved sequences misclassified by Kraken2. The reverse comparison (organisms reported by DME+ but not recovered by Kraken2/Bracken) was also examined. These calls were not subjected to the same BLASTn validation, as DME+ (the targeted, threshold-based assay) served as the reference standard for these organisms.

## Results

### Development of a Rapid Hybrid Capture Workflow (Rapid-RPIP v1)

To develop and optimise a rapid hybrid capture workflow, we systematically refined multiple steps of the RPIP protocol by evaluating alternative methods that offered workflow simplification or shorter processing times. The resulting workflow (Rapid-RPIP v1) reduced the pre-sequencing processing time from approximately 11.4 to 7.3 h, representing a reduction of approximately 4.1 h (Figure 1A).

**Figure 1.**
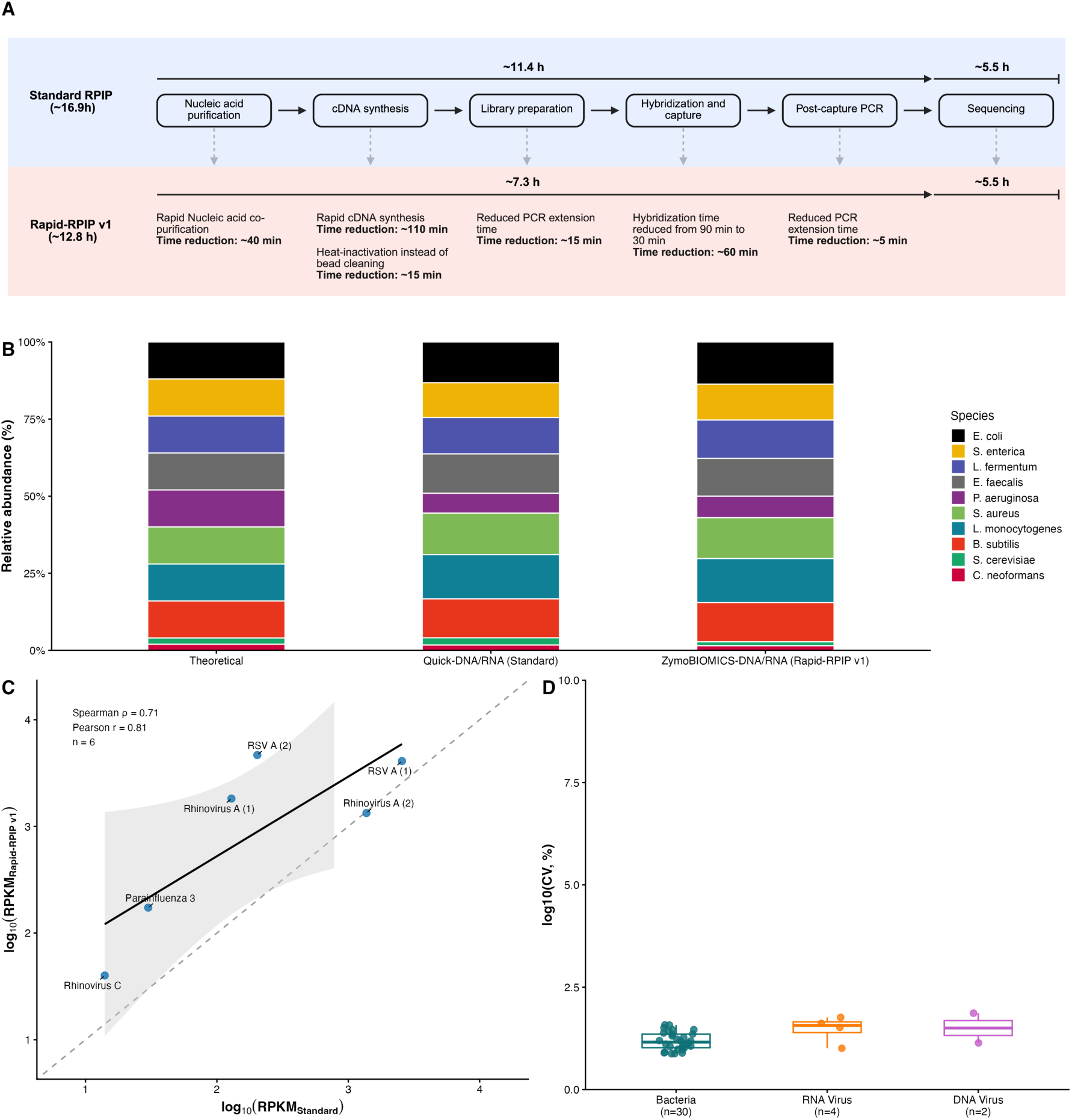
Wet-lab protocol optimisation and validation. (A) Schematic overview of the protocol modifications incorporated into Rapid-RPIP v1 relative to the Standard RPIP workflow. Individual optimisation steps, selected to reduce turnaround time, and their estimated time savings are shown. (B) Relative abundance (%) of the ten ZymoBIOMICS-DNA/RNA mock community organisms recovered by two extraction methods (Quick-DNA/RNA, ZymoBIOMICS-DNA/RNA; mean of 2 replicates each) compared with the manufacturer theoretical composition. (C) RPKM concordance between standard and rapid cDNA synthesis protocols for RNA respiratory viruses (n = 6 organism–sample pairs). Dashed line: identity (y = x); solid line: linear regression with 95% CI. Spearman ρ and Pearson r (log₁₀-RPKM) annotated. (D) Coefficient of variation (log₁₀ scale) of RPKM values across three hybridisation times (30/60/90 min), grouped by pathogen type (Bacteria, RNA Virus, DNA Virus); n per group annotated.

Focussing first on the nucleic acid extraction, we showed comparable recovery of the ZymoBIOMICS microbial community standards with the Quick-DNA/RNA Pathogen Miniprep Kit and the ZymoBIOMICS DNA/RNA Miniprep kit used in the Standard RPIP protocol (Figure 1B), reducing sample processing time by approximately 40 min (Figure 1A). We next demonstrated a saving of 1 h and 50 min by substituting the Standard RPIP cDNA synthesis workflow with the rapid cDNA synthesis protocol [14] . When tested on a panel of clinical samples, the latter also generated higher RPKM values for most RNA viruses (5 of 6 organism–sample pairs; median 4.3-fold higher, range 0.97-fold to 23.0-fold; Figure 1C). To optimise the hybridisation step, hybridisation times of 90 min (standard), 60 min, and 30 min were compared using libraries from samples positive for a range of DNA viruses, RNA viruses, and bacteria (Figure 1D). RPKM values for individual organisms remained broadly stable across hybridisation times, with a median coefficient of variation of 15.4% (range 7.5– 73.3%) across the 36 organism-sample measurements, and no association between variability and organism abundance (Spearman ρ = −0.04). Variability was comparable across pathogen types, though group sizes were unbalanced (bacteria, n = 30; RNA viruses, n = 4; DNA viruses, n = 2).

Finally, we incorporated several minor refinements, including replacing bead-based purification with heat inactivation following cDNA synthesis and optimising PCR conditions which further reduced workflow time by approximately 35 min. Taken together, all the above modifications comprised the Rapid-RPIP v1 workflow. When combined with the MiniSeq Rapid kit (∼5.5 h sequencing time) and automated downstream analysis (∼1 h), the complete workflow delivered results in less than 15 h from sample to final result.

### Evaluation of Rapid-RPIP v1 on mock community controls

Having optimised the components of a Rapid-RPIP v1, we evaluated the performance of the workflow compared to the Standard RPIP workflow using the NATtrol Respiratory Panel 2.1, which contains two controls, each one with a mix of DNA and RNA viruses and bacteria (Table 1). Each control was processed as supplied and at a 10-fold dilution. Analysis of undiluted Control 1 demonstrated successful detection of all eleven targeted pathogens using both sequencing protocols (Figure 2A). Adenovirus Type 31 is not targeted by the RPIP panel and is not part of the DME+ database, so it was not detected by either the Standard RPIP or Rapid-RPIP v1 protocols when analysis was performed using DME+. However, Kraken2/Bracken analysis of off-target reads detected Adenovirus Type 31 in both workflows. Analysis of Control 2 with DME+ detected ten targeted pathogens across both the Standard and Rapid-RPIP v1 protocols (Figure 2A). Coronavirus HKU-1, for which, unlike other pathogens, only a short recombinant sequence is present in the NATrol respiratory Panel, and not a full inactivated organism, was missed by both protocols using DME+. Kraken2/Bracken detected HKU-1 in the Rapid-RPIP v1 workflow only.

**Figure 2.**
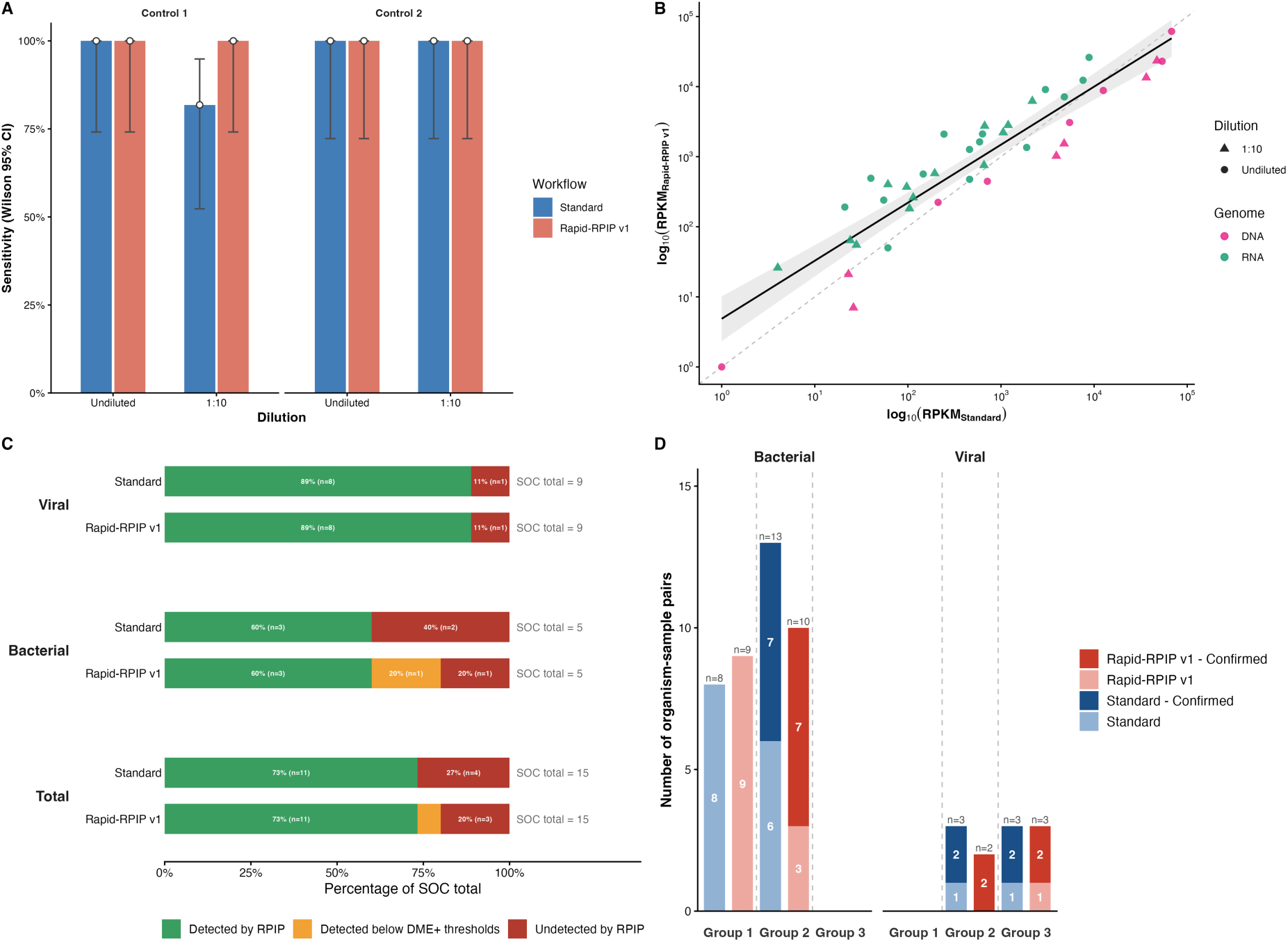
Comparison of Standard and Rapid-RPIP v1 protocols. (A) Sensitivity by dilution in mock community controls (Control 1 and Control 2; undiluted, 1:10), with Wilson 95% confidence intervals, by protocol. (B) RPKM concordance in mock community controls: log₁₀-transformed RPKM values for each organism detected by both protocols, coloured by nucleic acid type (RNA vs DNA), with dashed line indicating identity (y = x) and fitted regression. (C) Detection by the RPIP protocols of SOC-confirmed organisms in clinical samples, split by viral, bacterial and total categories, expressed as percentage of SOC total per protocol with n values annotated. Missed detections are further split into below-threshold detections (RPKM > 1 in DME+ below-thresholds and not called at the standard reporting threshold) versus genuine misses (false negatives). (D) Pathogens detected by both RPIP workflows but not by SOC in clinical samples, shown separately for bacterial and viral organisms and grouped by DME+ phenotypic classification (Group 1: commensal/low pathogenic potential organisms; Group 2: opportunistic or possible pathogens; Group 3: established respiratory pathogens), further stratified by confirmation status.

At 10-fold dilution, the two protocols diverged in Control 1 but not in Control 2 (Figure 2A). In Control 1, Standard RPIP failed to detect Influenza A H3N2 and HPIV-4, whereas Rapid-RPIP v1 detected both organisms. For Influenza A H3N2, Standard RPIP recovered 22775 aligned reads (RPKM = 505.5, 87.2% coverage) that nonetheless fell below the DME+ reporting threshold due to Average Nucleotide Identity (ANI; 90.5%, vs 92.1% for Rapid-RPIP v1), suggesting a reference matching effect rather than a true absence.

Kraken2/Bracken also detected parainfluenza virus reads in Control 1 in both protocols, consistent with a dilution-sensitive detection rather than an artifact. In Control 2, both protocols detected all 10 expected organisms (Table S2).

Although both protocols achieved similar qualitative detection, RPKM values differed systematically. In undiluted controls the two methods were strongly concordant for RPKM (Spearman rho = 0.92, p = 5.8X10^-26^; Pearson r on log₁₀-RPKM = 0.91, p = 1.9x10^-24^) for 41 pathogens detected by both workflows. RPKM concordance remained consistent for both undiluted and diluted samples (Spearman rho = 0.90 undiluted, n=22; 0.93 at 1:10, n=19), confirming that relative read-abundance was preserved between Standard RPIP and Rapid-RPIP v1 even when the input was reduced. Although concordance was high overall, the relative RPKM values for the two protocols differed between DNA and RNA pathogens (Figure 2B). RNA pathogens tended to give higher RPKM with the Rapid-RPIP v1 workflow (median log₂(Rapid-RPIP v1/Standard) of +1.49 [IQR +0.93 to +1.97], n = 28), whereas DNA pathogens tended to give lower RPKM (median log₂(Rapid-RPIP v1/Standard) of −0.83 [IQR −1.43 to −0.15], n = 13; Mann-Whitney p = 1.1x10^-8^).

### Performance of Rapid-RPIP v1 on clinical samples

Twelve respiratory samples were processed through both workflows and compared with SOC testing (Table S3). Against routine SOC testing, both the Standard RPIP and Rapid-RPIPv1 protocols achieved equivalent overall sensitivity of 73.3% (Figure 2C; 11/15). Sensitivity was also identical between the protocols for viruses (89% i.e. 8/9) and bacteria (60% i.e. 3/5). Fisher’s exact test showed no significant difference in sensitivity between protocols (p = 1.0), confirming that the rapid workflow did not compromise diagnostic accuracy relative to the standard procedure.

Four pathogens detected by SOC were missed by both RPIP protocols (Figure 2C). *Mycoplasma pneumoniae* and Rhinovirus A (both sample SR-01) showed no reads in either protocol even when DME+ thresholds were removed. Both organisms however were reliably detected by DME+ in other samples: *Mycoplasma pneumoniae* at RPKM up to 714 in RP2.1 mock community controls, and Rhinovirus A family members were detected in 21 other clinical samples, indicating that the panel and reference database can detect both organisms. Thus, their absence in sample SR-01 reflects a sample-specific failure rather than a systematic detection gap. This raises the possibility of sample degradation as a contributing factor, potentially due to storage at +4°C for more than 24 h before DNA/RNA Shield was added. In contrast, *Candida albicans* (SR-06) was absent from the DME+ output even when the thresholds were removed and was not detected by DME+ in any sample across the study. This is expected, as it is neither targeted by RPIP nor present in the DME+ database. The fourth pathogen, *Stenotrophomonas sp.* (SR-06), was a genuine below-threshold detection rather than a true absence, falling below the DME+ reporting threshold in Rapid-RPIP v1 (RPKM = 4.7) while remaining undetected by the Standard protocol.

Both Standard and Rapid-RPIP v1 workflows identified organisms not detected by SOC, that were independently confirmed by additional testing (Figure 2D), including RNA viruses (Influenza B virus, Rhinovirus A, Rhinovirus C and Coronavirus OC43), DNA viruses (Human adenovirus C) and bacteria (*Streptococcus pneumoniae, Pseudomonas aeruginosa* and *Haemophilus influenza*e).Figure 2D also includes organisms detected only by both RPIP workflows and not by SOC, which have not been independently tested by any additional method (Table S4). In two cases of Human adenovirus C detection, the RPKM in the negative control from the same sequencing batch were similar (RPKM 2.3–2.8 in negative control vs. 5.1–8.5 in sample) suggesting that contamination was the likely cause. A third Human Adenovirus C could not be explained by contamination and was considered as a true positive. Additional targeted bacteria were detected but not confirmed by other methods; these fell into Groups 1 and 2 i.e. commensals that can occasionally be pathogenic and showed lower RPKM than confirmed detections (median 5.4 vs. 28.8), consistent with low-level colonisation rather than confirmed infection (Figure 2D).

As with the mock communities, in five of six cases, RNA viruses had higher RPKM in the Rapid-RPIP v1 protocol (median 1.95-fold higher, +95%; range 0.51-fold to 10.2-fold). This was most pronounced for Human metapneumovirus (Rapid-RPIP v1: 2,802 vs Standard: 541 RPKM; 5.2-fold) and Human respiratory syncytial virus B (Rapid-RPIP v1: 474 vs Standard: 47 RPKM; 10.2-fold). The two protocols were strongly concordant on the log scale (n = 34 paired observations; Spearman ρ = 0.91, p <2×10⁻¹⁶; Pearson r on log₁₀-RPKM = 0.93, p = 3.5×10⁻¹⁵). Again, similarly to the mock samples, DNA targets showed lower RPKM for Rapid-RPIP v1 compared with Standard RPIP in 68% of paired results (median 21% lower, n=28). Human cytomegalovirus and Epstein-Barr virus were detected by both protocols although with lower abundance in Rapid-RPIP v1 (CMV: 554 vs 311 RPKM, 44% lower; EBV: 11.1 vs 2.5 RPKM, 78% lower), consistent with the reduced numbers of DNA reads observed in mock communities (Figure S1). *Alternaria infectoria* was detected in one sample (SR-08) with markedly lower abundance in the Rapid-RPIP v1 protocol (Rapid-RPIP v1: 0.77 vs Standard: 35.51 RPKM); this organism was not part of SOC testing and was not independently confirmed, representing a notable discrepancy between workflows that warrants further investigation.

### Development of an Updated Rapid Hybrid Capture Workflow (Rapid-RPIP v2)

Following validation of the initial workflow, we sought to further reduce turnaround time and simplify the workflow complexity. Illumina updated the Respiratory Pathogen Identification/AMR Enrichment Panel kit to incorporate an integrated approach combining cDNA synthesis and bead-linked transposase (BLT) fragmentation into a single reaction, which we found reduced library preparation time by approximately 25 min compared to the Rapid-RPIP v1 protocol. We implemented an updated version of the protocol (Rapid-RPIP v2) by incorporating all other modifications from Rapid-RPIP v1 into the updated kit.

In parallel, sequencing was transitioned to the MiSeq i100 platform using a Turbo Sequencing program, which simplifies loading and reduces run time by approximately 3 h compared to the Rapid MiniSeq sequencing kit and by approximately 2 h compared to the standard MiSeq i100 protocol. Collectively, these improvements reduced the wet-lab workflow to approximately 6.6 h and enabled a total sample-to-result turnaround time of under 10 h (Figure 3A).

**Figure 3.**
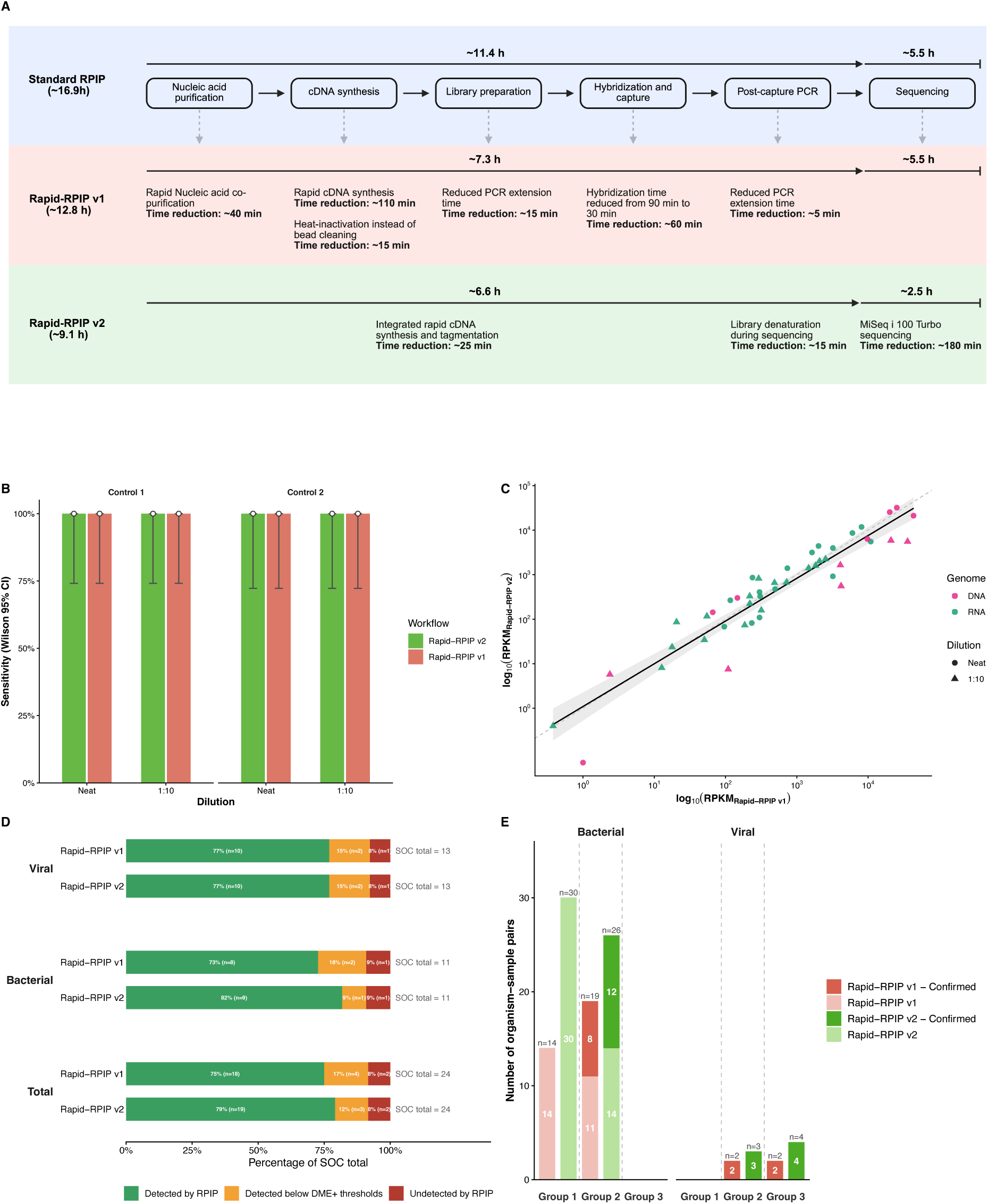
Comparison of Rapid-RPIP v1 and Rapid-RPIP v2 protocols. (A) Schematic overview of the protocol modifications incorporated into Rapid-RPIP v2 relative to the Standard RPIP and Rapid-RPIP v1 workflows. Individual optimisation steps, selected to reduce turnaround time, and their estimated time savings are shown. (B) Sensitivity by dilution in mock community controls (Control 1 and Control 2; undiluted, 1:10), with Wilson 95% confidence intervals, by protocol. (C) RPKM concordance in mock community: log₁₀-transformed RPKM values for each organism detected by both protocols, coloured by nucleic acid type (RNA, DNA), with dashed identity line (x=y) and fitted regression. (D) Detection by the RPIP workflows of SOC-confirmed organisms in clinical samples, split by viral, bacterial and total categories, expressed as percentage of SOC total per protocol. Missed detections are further split into below-threshold detections (RPKM > 1 in the below-threshold DME+ export but not called at the standard reporting threshold) versus genuine misses. (E) Pathogens detected by both RPIP workflows but not by SOC in clinical samples, shown separately for bacterial and viral organisms and grouped by DME+ phenotypic classification (Group 1: commensal/low pathogenic potential organisms; Group 2: opportunistic or possible pathogens; Group 3: established respiratory pathogens), further stratified by confirmation status.

### Evaluation of Rapid-RPIP v2 on mock community controls

To evaluate the performance of the Rapid-RPIP v2 workflow compared to Rapid-RPIP v1, we used the same approach as for comparing Rapid-RPIP v1 and Standard RPIP. The NATtrol Respiratory Panel 2.1. was processed as supplied and at a 10-fold dilution. Both workflows achieved 100% sensitivity for targeted organisms in undiluted and 10-fold diluted samples across both control panels (Figure 3B; Control 1: 11/11; Control 2: 10/10 at both dilution levels). Unlike the Standard vs Rapid-RPIP v1 comparison, where Kraken2/Bracken detected HKU-1 in the off-target reads, no HKU-1 signal was found by Kraken2/Bracken in either protocol.

The two protocols were strongly concordant across all pathogens in the mock community detected by both Rapid-RPIP v1 and Rapid-RPIP v2 (n = 46 paired observations; Figure 3C; Spearman ρ = 0.94, p < 2.2×10⁻¹⁶; Pearson r on log₁₀-RPKM = 0.94, p = 6.6×10⁻²²). This indicates that relative organism abundance was preserved between workflows (Table S5). DNA organisms tended to give lower RPKM with Rapid-RPIP v2 vs Rapid-RPIP v1 (median 51% lower; IQR: 85% lower to 27% higher; n = 13), while RNA organisms were broadly similar between Rapid-RPIP v1 and Rapid-RPIP v2 (median 5% higher; IQR: 29% lower to 51% higher; n = 33), though this difference between the RNA and DNA fold change distributions did not reach statistical significance (Mann-Whitney p = 0.063).

### Performance of Rapid-RPIP v2 on clinical samples

Twenty-three respiratory samples were processed with both Rapid-RPIP v1 and Rapid-RPIP v2 and compared against routine SOC testing in a separate clinical cohort (Table S6). Across the 24 organisms confirmed positive by SOC testing (some samples had more than one confirmed pathogen, each counted separately), Rapid-RPIP v2 achieved sensitivity of 79.2% (19/24), compared to 75% (18/24) for Rapid-RPIP v1 (Fisher’s exact test, p = 1). Each protocol uniquely detected one organism that the other protocol detected below the threshold rather than missing: Rapid-RPIP v2 detected *Pseudomonas aeruginosa* (RT-13), which Rapid-RPIP v1 recovered below the DME+ threshold (RPKM = 9.9, ANI = 99.1%, 580 aligned reads, 20.6% coverage), while Rapid-RPIP v1 detected Rhinovirus A (RT-20; RPKM = 44,993), which Rapid-RPIP v2 also recovered but below threshold despite greater read depth (RPKM = 67,644, ANI = 93.7%, 1,035,515 aligned reads, 99.9% coverage).

Among the pathogens detected by SOC but missed by both Rapid-RPIP v1 and Rapid-RPIP v2 protocols (Figure 3D), two reflected genuine absence even when thresholds were not considered: *Enterobacter hormaechei* and Adenovirus. A third pathogen, *Staphylococcus aureus* (in the same sample as the *Pseudomonas aeruginosa* case above, RT-13), was detected at levels below the DME+ threshold for calling a positive by both protocols (Rapid-RPIP v1: RPKM = 3.3; Rapid-RPIP v2: RPKM = 1.3).

Influenza A virus was detected in three clinical samples (RT-17: H1N1; RT-18 and RT-19: H3N2). Both the Rapid-RPIP v1 and Rapid-RPIP v2 protocols reconstructed near-complete genomes across all eight influenza gene segments for all three samples (mean per-segment consensus coverage >98.5% for both Rapid-RPIP v1 and v2), with concordant subtype calls between protocols in every case. Segment-level coverage maps for these samples are provided in Figure S2.

We also examined RPKM values between Rapid-RPIP v1 and Rapid-RPIP v2 across all the organisms detected by either protocol in clinical samples (Figure S3). The Rapid-RPIP v1 and Rapid-RPIP v2 workflows were strongly concordant on the log scale (n = 49 paired observations; Spearman ρ = 0.80, p < 2×10⁻¹⁶; Pearson r on log₁₀-RPKM = 0.85, p = 2.3×10⁻¹⁴). Consistent with the mock community comparison (Figure 3C), RNA and DNA differences did not reach statistical significance in clinical samples either (RNA median log₂(Rapid v2/Rapid v1) = +0.63, n = 11; DNA median −0.17, n = 38; Mann-Whitney p = 0.34).

Beyond the SOC-confirmed pathogens, both protocols identified additional organisms spanning a wide RPKM range (RPKM 0.15–10,642; median 74.7 across both protocols) which included respiratory bacteria and viruses not captured by SOC but confirmed by additional testing. We were unable to identify Ct or CFU levels associated with detection or not of an organism as these data were not available. Organisms detected exclusively by both Rapid-RPIP workflows are summarised in Figure 3E. Additional bacteria were detected by the Rapid-RPIP workflows but not confirmed by other methods; these fell into Groups 1 and 2, i.e. likely to be commensal organisms, and showed lower RPKM than confirmed detections (median 20.3 vs. 71), consistent with low-level colonization rather than confirmed infection (Table S4).

### Secondary analysis to confirm detection

To investigate organisms missed by DME+, we used Kraken2/Bracken for taxonomic classification. This identified three organisms (Cytomegalovirus, Enterovirus, and *Pseudomonas aeruginosa*) that DME+ missed in some samples, even though they are in the DME+ database and were detected in other samples from this cohort.

Cytomegalovirus (RT-13) reads matched Human betaherpesvirus 5 at 99–100% identity; this finding was confirmed by confirmatory PCR, with the organism present at RPKM 0.7–1.3 (57–59 aligned reads), below the DME+ threshold. Enterovirus (RT-10) reads matched Coxsackievirus B4 at 98–100% identity across 9/10 reads (with one read matching Enterovirus B); this finding was confirmed by routine SOC testing, with reads distributed across multiple related serotypes, each individually below threshold. *Pseudomonas aeruginosa* (RT-13) reads matched P*. aeruginosa* reference strains at 100% identity across all 10 reads; this finding was confirmed by routine SOC testing, with the organism present at RPKM 7.2–9.9 (417–580 aligned reads), below the DME+ threshold. In the same sample as this *Pseudomonas aeruginosa* case (RT-13), the single false positive was *Staphylococcus aureus*: all 10 Kraken2 classified reads mapped to *Pseudomonas aeruginosa* at reduced identity (91–92%, alignment length 66–74 bp), indicating that conserved *Pseudomonas* sequences were misclassified by Kraken2 as *Staphylococcus aureus*.

An additional finding, *Candida albicans* reads (sample RT-15) matched *C. albicans* database sequences at 95–100% identity, confirming its presence despite the DME+ call being negative; this finding was also independently verified by confirmatory PCR. This is consistent with *Candida albicans* not being targeted by the panel and therefore not represented in the DME+ reference database, rather than a true absence. Mapping of Kraken2 classified reads to reference genomes showed sparse coverage for all five Kraken2-positive/DME+-negative discordant organisms (<3% genome coverage, <0.5× depth) insufficient to distinguish low-abundance from classification noise. BLASTn validation of the Kraken2 classified reads against the NCBI nr/nt database was therefore performed for all five (see Methods).

To assess whether background contamination could account for any reported detections, we compared organism detected in clinical samples against ten sequenced negative controls (NC1, NC3–NC11; NC2 was excluded from analysis due to an indexing error during library preparation; multiple negative controls (NCs) were included per run, so this did not impact result interpretation) processed across the study. In the DME+ output, the NCs were clean for every organism under scrutiny with one exception: Rhinovirus A showed low-level background in two of ten controls (RPKM 1.7 and 95.0); both belonged to sequencing batches unrelated to the sample in which Rhinovirus A was detected (the NCs included in the same batch showed no Rhinovirus A reads).

With Kraken2/Bracken, *Staphylococcus aureus* was present at high abundance in all ten NCs (30,222–283,689 reads), providing independent support for its classification as a false positive rather than a genuine detection (see Secondary analysis, above). *Pseudomonas aeruginosa* and *Stenotrophomonas maltophilia* showed low-level background across all ten NCs (44–1,148 and 32–1,344 reads, respectively), whereas *Candida albicans*,

Cytomegalovirus and Enterovirus were entirely absent from every NC, supporting these as genuine detections.

## Discussion

We have developed and evaluated a same-day hybrid capture workflow, Rapid-RPIP v2, for the detection of respiratory pathogens based on Ilumina’s RPIP panel. Our workflow reduced the sample processing wet-lab time from around 11.4 to approximately 6.6 h without compromising sensitivity. When combined with rapid sequencing programs, like Turbo sequencing for the MiSeq i100, and automated analysis, the workflow provides results in under 10 h.

Both the Rapid-RPIP v1 and Rapid-RPIP v2 workflows achieved higher sensitivity than the Standard workflow across mock communities with strong quantitative concordance throughout (Spearman ρ=0.90–0.94). In clinical samples, sensitivity was statistically equivalent across all comparisons (Standard vs Rapid-RPIP v1: both 73.3%, Fisher p=1.0; Rapid-RPIP v1 vs Rapid-RPIP v2: 75.0% vs 79.2%, Fisher p=1). Overall, performance of RPIP workflows in clinical samples was slightly better for viruses compared to bacteria (Figure 2C, 3D). Bacterial sensitivity improved between Rapid-RPIP v1 and v2 specifically (73% to 82%), though it did not change between the Standard and Rapid-RPIP v1 workflows (60% for both); further analyses of a larger cohort are required to determine whether this reflects a protocol improvement or cohort specific variation.

Where pathogens detected by SOC were missed by RPIP, causes fell into three categories. First, one sample (SR-01) showed no reads for either of two SOC-confirmed organisms (*Mycoplasma pneumoniae* and Rhinovirus A) in any protocol. This together with detection of these organisms in other samples, raises the possibility of sample degradation as a cause. The sample in question had been kept at +4^0^C for two days before the preservation solution DNA/RNA Shield was added and the sample frozen. Second, the presence of organism reads but at levels below the DME+ threshold for reporting a positive result. For example, *Stenotrophomonas sp.* was recovered below threshold by Rapid-RPIP v1 (RPKM = 4.7) after being missed by Standard RPIP, and in the Rapid-RPIP v1 and v2 comparison, *Pseudomonas aeruginosa*, *Staphylococcus aureus* and Rhinovirus A were each recovered below-threshold in at least one protocol. Third, where missed detections, even when DME+ thresholds were removed. In these cases, the reasons differed by organism. *Enterobacter hormaechei* (RT-04) had no reads in either Rapid-RPIP protocol, consistent with a probe-capture failure rather than a downstream classification issue. In contrast, *Candida albicans* (SR-06) was negative on DME+, even in the absence of thresholds and when analysed using Kraken2/Bracken. In another clinical sample (RT-15), it was detected at low depth and coverage by Kraken2/Bracken and confirmed by BLASTn. As RPIP targets specific regions of *Candida auris* rather than *Candida albicans*, the detection in RT-15 likely reflects detection of off-target reads rather than true probe-based enrichment.

Beyond SOC-confirmed pathogens, both Rapid-RPIP v1 and Rapid-RPIP v2 protocols consistently identified additional organisms which were subsequently independently confirmed by additional testing, including respiratory viruses, herpesviruses, and bacterial pathogens across both clinical cohorts (Figure 2D, Figure 3E), demonstrating that neither rapid workflow compromised the broader detection capability of the RPIP panel relative to the standard protocol. This is clinically relevant given that SOC testing typically detects only culturable organisms plus a limited, predefined pathogen set that can be detected by PCR, whereas hybrid capture can recover a broader range of clinically significant organisms without additional pre-specified testing. The sensitivity of hybrid capture approaches is highly reliant on the quality of and diversity in the panel design. Large comprehensive panels are commercially available for virus detection and provide whole genome sequence information [17,20]. While this is feasible for viruses, given their comparatively small genome sizes, one limitation of the RPIP panel, particularly for bacteria and fungi, is that only small regions of the genome that support identification and/or provide resistance information are targeted [19,21]. This is a common constraint of hybrid capture methods, since targeting full bacterial and fungal genomes would require probe panels of substantially greater size and cost. Expansion of the RPIP probe set to include a broader range of pathogens and increased coverage of existing targets could further improve pathogen detection and facilitate downstream genomic analyses.

Implementation of an automated data analysis pipeline, such as DME+, with predetermined detection thresholds reduces the need for complex bioinformatic analyses and facilitates interpretation of metagenomic results. Furthermore, DME+ assigns positive organisms to phenotypic Groups 1, 2, or 3, providing additional context for result interpretation. This is particularly valuable for non-sterile sites, such as respiratory samples, where resident microorganisms are expected to be detected. When considered alongside RPKM values, this classification can help assess the clinical relevance of detected organisms and distinguish potential pathogens from background microbial flora.

One of the limitations of DME+ is that its database only contains the targeted regions of the species in the panel, meaning that information provided by off-target reads and non-targeted related organisms will be missed. Currently, complementary analysis with other classifiers and larger databases may be needed to recover this additional information. Four low-abundance organisms negative by DME+: *Candida albicans* and Cytomegalovirus (both confirmed by PCR), Enterovirus, and *Pseudomonas aeruginosa (*detected by SOC) were nonetheless identified using Kraken2/Bracken and confirmed as genuine detections by BLASTn. For a fifth organism*, Staphylococcus aureus* (also SOC positive), only a small number of Kraken2 classified reads were recovered and BLASTn showed these matched to *Pseudomonas aeruginosa* reference sequences at reduced identity (91-92%) rather than confirming *Staphylococcus aureus*. This illustrates the limits of both RPIP and k-mer-based classifiers [32]: at low abundance, signal can be swamped by more abundant co-occurring pathogens, and because probes only target small regions of the genome, the reads recovered may not be sufficiently discriminatory to confirm the initial call. Notably, genome coverage metrics alone were insufficient to resolve these calls, particularly for bacterial and fungi targets, where information on the regions targeted by the RPIP panel is not available.

In addition to database limitations, the presence of predefined thresholds in DME+, particularly in cases where metrics such as coverage and ANI are included, can make analysis challenging. This is illustrated by sample RT-23, where both protocols recovered Rhinovirus A with high coverage and very high RPKM (Rapid-RPIP v1: RPKM 9,398, 83% coverage, 66,946 aligned reads; Rapid-RPIP v2: RPKM 137,004, 82% coverage, 483,660 aligned reads), yet both still failed the ANI threshold (92.5% and 91.9%, respectively). A similar pattern was seen in sample RT-20 for Rhinovirus A, where Rapid-RPIP v2 recovered substantially greater depth and coverage than v1 (RPKM = 67,644 vs. 44,993) yet still fell below threshold (ANI 93.7%). Together, these cases show that the effect is not systematically biased toward either protocol. The magnitude of read recovery makes clear that the limitation lies not in sequencing depth or capture efficiency, but in the ANI threshold itself. For highly diverse pathogens with limited reference database representation, a high ANI threshold can result in false-negative calls even when reads are recovered at high depth and coverage.

Another disadvantage of DME+ is that its analysis does not consider negative controls run in parallel with the samples being tested. This makes identification of contaminants introduced during sample preparation more challenging [3,4]. To address this, we added our own negative controls and compared results from samples and controls [19]. From this we were able to identify two instances of Adenovirus C contamination which were excluded from further analysis and to independently confirm the false-positive *Staphylococcus aureus* results described above.

Both Rapid-RPIP workflows (v1 and v2) showed an increase sensitivity for RNA targets compared with the Standard protocol (higher RPKM values; up to 181% higher in mock communities and 95% higher in clinical samples for Rapid-RPIP v1 vs Standard). Rapid-RPIP v2 was not directly compared with the Standard protocol; however, in the comparison between Rapid-RPIP v1 and Rapid-RPIP v2, the latter showed a modest increase in RPKM values over Rapid-RPIP v1 (Figure 3). Increased sensitivity for RNA targets, can help overcome the limitations of existing rapid untargeted metagenomic workflows, where a reduced sensitivity for RNA viruses like Influenza and Rhinovirus A has been observed [10,23]. One explanation for this improvement is the simplification of the cDNA synthesis step (fewer steps, shorter incubation times) and the elimination of a post-cDNA synthesis washing step. While the increase in RPKM for RNA targets lead to a decrease in RPKM for DNA targets (approximately 44% lower, consistent across both comparisons), this did not affect the sensitivity of detection of DNA pathogens in either rapid workflow.

An advantage of the approach we have taken here is that hybrid capture workflows can be modular such that individual steps can be optimised independently and new developments can be easily integrated into the protocol. Our aim was to reduce protocol complexity and turnaround times, without compromising workflow performance. Combining this approach with iterative method improvement (Rapid-RPIP v1, Rapid-RPIP v2) provides the basis for the translation of research-focused workflows to clinical settings. Evaluation of the current workflow (v2) on a larger sample set will help identify additional areas of improvement, for example, supplementing the panels with additional targets of interest, or assessing potential nucleic acid purification biases that may impact the detection of some organisms like fungi.

Finally, this workflow has the potential to be used in other scenarios, like central nervous system infections and sepsis. This can be achieved by adapting the nucleic acid purification to specific sample types (CSF, plasma, tissue, urine) and developing syndromic panels targeting pathogens associated with each scenario. While different metagenomic approaches have been developed and implemented for these use cases, they have limitations in terms of sensitivity, workflow complexity and long turnaround times [5,6,19]. Additionally, since hybrid capture workflows do not rely on host removal approaches to increase sensitivity, they can allow the analysis of host information, like host transcripts for assessment of infection-type and prognosis and the identification of germline and somatic mutations [19]. The development of a rapid integrated approach that provides information not only on the pathogen but also on the individual, provides the foundation for precision medicine approaches to diagnosis and patient management.

This study describes the development and validation of two rapid hybridisation capture workflows (Rapid-RPIP v1 and Rapid-RPIP v2), both substantially reducing sample-to-result turnaround time compared to the Standard RPIP protocol while maintaining clinical sensitivity against SOC-confirmed respiratory pathogens. High quantitative concordance was observed across the full pathogen abundance range for both comparisons, supporting the clinical viability of same-day hybrid capture workflows for respiratory infection diagnosis. Further studies using larger cohorts are needed to determine whether the reduced read recovery observed for DNA pathogens in the Rapid-RPIP protocols has clinical significance, and modifications to probe design and analysis pipeline thresholds may further improve sensitivity for low-abundance and poorly represented organisms.

We acknowledge several limitations. Both clinical validation cohorts were small, yielding wide confidence intervals that limit the precision of sensitivity estimates and preclude definitive conclusions about protocol differences. The two cohorts were recruited and processed separately, preventing direct three-way protocol comparison on the same samples. SOC testing was used as the reference standard, which has imperfect sensitivity for certain organisms and may contribute to apparent missed detections. We did not formally assess the limit of detection (LOD) of RPIP. This was partly because the NATtrol/Zeptometrix mock community controls are not quantitative, so we instead used a 10-fold dilution to provide some insight into sensitivity at reduced input. Similarly, quantitative measures of the original clinical specimen (Ct values for PCR-positive organisms, or CFU counts for culture-positive organisms) were not systematically available for this cohort, so we are unable to determine whether the pathogens missed by RPIP (*Mycoplasma pneumoniae*, *Candida albicans*, Rhinovirus A, *Enterobacter hormaechei* and Adenovirus, plus the below-threshold cases of *Stenotrophomonas sp*., *Pseudomonas aeruginosa* and *Staphylococcus aureus*) reflected genuinely low organism burden in the original sample or a limitation of the sequencing/bioinformatic workflow. A formal LOD assessment for RPIP, along the lines of those recently reported for other rapid respiratory metagenomic protocols [24,33], is needed and is the subject of a larger analysis currently underway. Findings from the Kraken2/Bracken analysis are exploratory and based on a small number of discordant calls; larger prospective studies would be needed to establish the added diagnostic value of parallel k-mer classification in routine practice. Bioinformatic analysis with the DME+ is also a limitation. The DME+ database is missing some organisms (e.g. *Candida albicans*), applies high detection thresholds, fails to take advantage of off-target reads and does not incorporate comparison with negative controls run in parallel. We have shown both here and in previous analyses [17] that incorporating each of these improves the accuracy of metagenomics reporting. Both rapid protocols achieved sensitivity and quantitative concordance equivalent to the standard workflow. Rapid-RPIP v2 showed a modest numerical performance advantage and represents the preferred candidate for wider clinical evaluation. A systematic RNA/DNA abundance bias was identified that does not compromise detection but should be accounted for in quantitative applications. While parallel Kraken2/Bracken classification provides complementary detection at the margins of DME+, with BLASTn confirmation required to resolve discordant calls in targeted sequencing data, better programmes that incorporate off-target read analysis, negative control comparison, and broader reference databases would improve overall reporting accuracy.

## Conclusions

We have developed a same-day hybrid capture workflow for respiratory pathogen detection that demonstrated performance comparable to SOC testing while identifying additional pathogens missed by routine clinical diagnostics. Our method enabled rapid recovery of full viral genomes, providing information beyond pathogen identification. Together, these capabilities highlight the potential of this approach to become a single-assay solution for both diagnosis and surveillance, enhancing patient management and informing public health responses. Given the modular nature of hybrid capture, this framework can be readily extended to other infectious disease and clinical applications.

## Supporting information

Rapid-RPIP v1 protocol updated

Rapid-RPIP v2 protocol updated

Clinical respiratory samples included in each protocol comparison

Detection performance of respiratory pathogens using standard and rapid sequencing protocols across undiluted and diluted samples

Detection and RPKM summary for ground-truth-positive pathogens, Standard vs Rapid-RPIP V1

Additional organisms detected beyond routine standard-of-care (SOC) testing, Figure 2D and Figure 3E

Detection performance of respiratory pathogens using Rapid-RPIP v1 and Rapid-RPIP v2 protocols across undiluted and diluted samples

Detection and RPKM summary for all ground-truth-positive pathogens, Rapid-RPIP v1 vs Rapid-RPIP v2

## List of abbreviations

AMR: Antimicrobial Resistance
ANI: Average Nucleotide Identity
BLASTn: Basic Local Alignment Search Tool (nucleotide)
DME+: DRAGEN Microbial Enrichment Plus v1.1.1
ES: Endo-tracheal Secretion
IQR: Interquartile Range
LOD: Limit of Detection
NPA: Nasopharyngeal Aspirate
RPIP: Respiratory Pathogen ID/AMR Enrichment Panel
RPKM: Reads Per Kilobase Million
RTI: Respiratory tract infections
SOC: Standard-of-Care testing

## Declarations

### Ethics approval and consent to participate

Residual clinical samples used for method development and validation were obtained from the UCL Pathogen Bank under ethical approval (REC reference: 12/LO/1089).

## Consent for publication

Not applicable

## Availability of data and materials

Raw sequencing reads are available from National Center for Biotechnology Information’s Sequence Read Archive under BioProject ID PRJNA1533207 (data will be made available after publication).

## Authors’ contributions

The study was designed by OETM, SM, JB, RW. Iterative workflow development was performed by OETM, RW, LMMB and TB. Clinical sample collection and sequencing was performed by TB, AK, GK, AL and LA. Clinical data collection was performed by LA, AL and JRB. Data curation was performed by: LB, AB, SB, SM. Bioinformatic and Statistical analysis: LB, AB, SB, SM. LB, JB, SM and OETM wrote and revised the manuscript. JRB, PS, HH, TG, RW and SC revised the manuscript. All authors read and approved the final manuscript.

## Acknowledgements

All samples in this study were processed and sequenced at UCL Genomics (RRID:SCR_027010)

## Supplementary Tables

**Table S1. Clinical respiratory samples included in each protocol comparison.** Each row is one clinical respiratory specimen processed through both protocols of the comparison indicated (Standard vs Rapid-RPIP v1, n = 12; Rapid-RPIP v1 vs Rapid-RPIP v2, n = 23). Column “sample_type” indicates the specimen type: NPA, nasopharyngeal aspirate; ES, endo-tracheal secretion. “pathogens_detected” lists the pathogen(s) confirmed by routine standard-of-care (SOC) testing for that sample. The entry “None (routine-SOC-negative)” indicates a sample with no routine-SOC-confirmed pathogen. Enterovirus and Rhinovirus A are reported as a combined category (“Enterovirus/Rhinovirus”), consistent with the routine PCR assay, which does not distinguish them.

**Table S2: Detection performance of respiratory pathogens using standard and rapid sequencing protocols across undiluted and diluted samples.** Respiratory pathogens present in the RP2.1 Control 1 and Control 2 mixtures were analysed using two distinct sequencing methodologies (standard and rapid protocols) at undiluted and 10-fold diluted concentrations. Detection results are presented as reads per kilobase per million mapped reads (RPKM), with numerical entries indicating successful pathogen identification and quantification. “ND” (not detected) indicates failure to detect the respective pathogen under the specified conditions.

**Table S3. Detection and RPKM summary for ground-truth-positive pathogens, Standard vs Rapid-RPIP V1.** One row per sample × pathogen × protocol combination, for pathogens confirmed by routine standard-of-care (SOC) testing: sample, clinical sample identifier; soc_pathogen, pathogen name; protocol, sequencing protocol (Standard or Rapid-RPIP v1); RPKM_sample, RPKM of the matched organism in the clinical sample (NA if not detected by DME+); RPKM_NC, maximum RPKM of a matching organism observed in any negative control run under the same protocol (0 if never called above threshold in any negative control).

**Table S4. Additional organisms detected beyond routine standard-of-care (SOC) testing, Figure 2D and Figure 3E**. One row per sample × organism × protocol combination for organisms detected by RPIP sequencing (DME+) that were not part of the routine SOC pathogen panel for that sample (i.e., not already covered by Tables S5–S6). Cohort indicates the protocol comparison (Standard vs Rapid-RPIP v1, corresponding to Figure 2D; Rapid-RPIP v1 vs Rapid-RPIP v2, corresponding to Figure 3E); sample, short sample identifier (as in Table 1); Protocol, sequencing protocol; Pathogen, organism detected; class_group, Bacterial or Viral; RPKM_Illumina, N_AlignedReads_Illumina and Coverage_Illumina, the RPKM, aligned read count, and genome coverage reported by DME+ for that organism in that sample/protocol; status indicates whether the finding was subsequently confirmed by an independent test (“Confirmed by another test”) or remains an RPIP-only, unconfirmed finding (“Illumina only (unconfirmed)”).

**Table S5: Detection performance of respiratory pathogens using Rapid-RPIP v1 and Rapid-RPIP v2 protocols across undiluted and diluted samples.** Respiratory pathogens present in the RP2.1 Control 1 and Control 2 mixtures were analysed using two distinct sequencing methodologies (Rapid-RPIP v1 and Rapid-RPIP v2) at undiluted and 10-fold diluted concentrations. Detection results are presented as reads per kilobase per million mapped reads (RPKM), with numerical entries indicating successful pathogen identification and quantification. “ND” (not detected) indicates failure to detect the respective pathogen under the specified conditions.

**Table S6. Detection and RPKM summary for all ground-truth-positive pathogens, Rapid-RPIP v1 vs Rapid-RPIP v2.** One row per sample × pathogen × protocol combination, for pathogens confirmed by routine standard-of-care (SOC) testing: sample, cinical sample identifier; soc_pathogen, pathogen name; protocol, sequencing protocol (Rapid-RPIP v1 and Rapid-RPIP v2); RPKM_sample, RPKM of the matched organism in the clinical sample (NA if not detected by DME+); RPKM_NC, maximum RPKM of a matching organism observed in any negative control run under the same protocol (0 if never called above threshold in any negative control).

**Table S7. BLASTn validation of Kraken2-classified reads from Bracken-discordant pathogen–sample pairs.** For each of the five organisms showing discordant results (Kraken2/Bracken positive, DME+ negative), the top ten Kraken2-classified reads were queried against the NCBI nr/nt database using BLASTn. Columns show the read identifier, top hit accession number and organism, percent nucleotide identity, alignment length (bp), E-value, and bitscore. Organisms confirmed at ≥95% identity to the expected species across the majority of reads were classified as genuine detections missed by DME+; reads mapping to an unexpected organism or at substantially reduced identity were classified as Kraken2 false positives.

## Supplementary Figures

**Figure S1.**
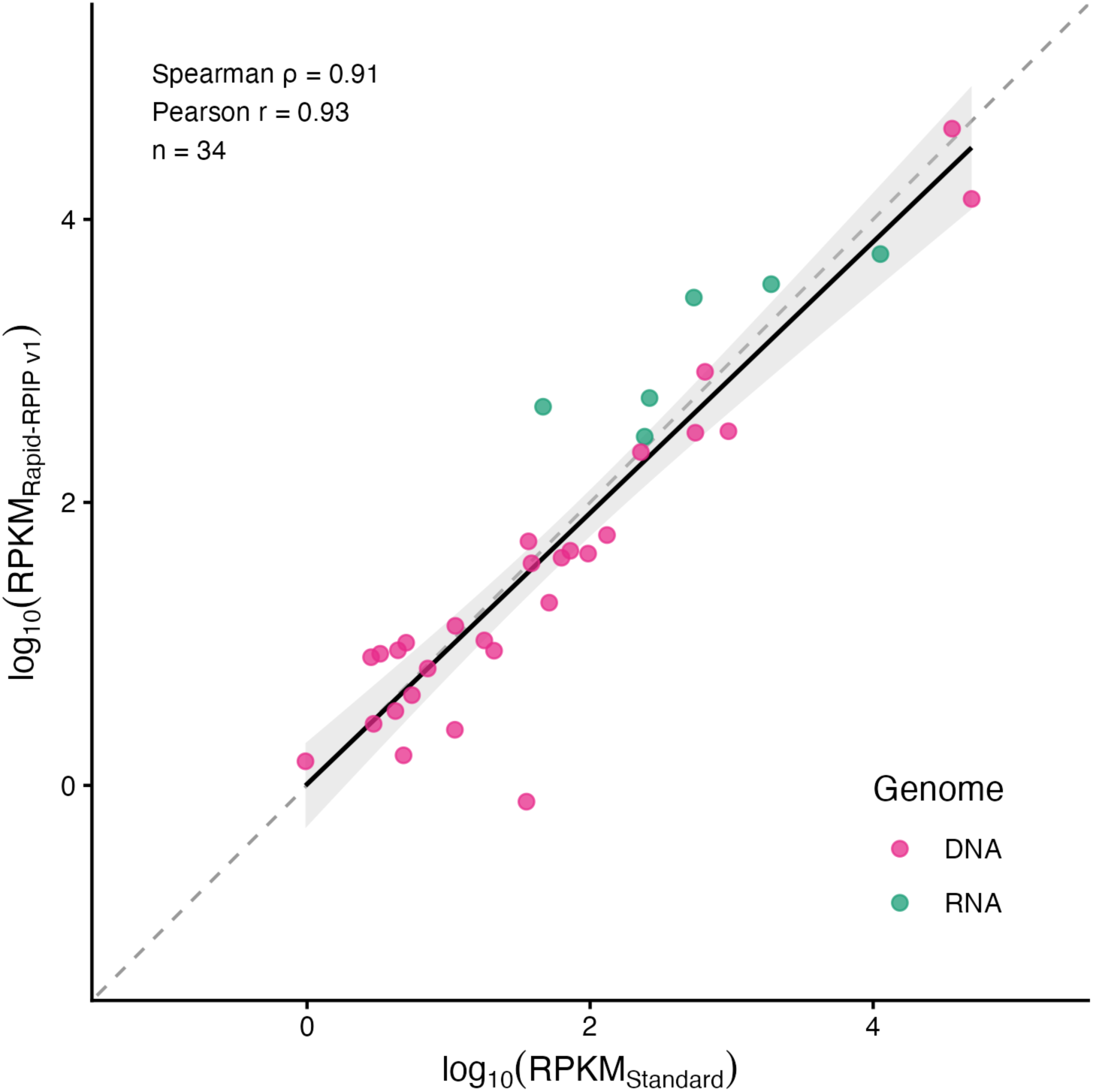
Quantitative concordance between Standard-RPIP and Rapid-RPIP v1 in clinical samples. Scatter plot of log₁₀-transformed RPKM values for organisms detected above threshold by both protocols in the same clinical sample (n=34 paired observations). Each point represents one organism sample pair, coloured by genome type (RNA, blue; DNA, red). The dashed grey line indicates the identity line (y=x); the solid black line with grey ribbon shows the linear regression with 95% confidence interval fitted on log₁₀-RPKM values. Spearman ρ was calculated on raw RPKM; Pearson r was calculated on log₁₀-RPKM. Organisms detected by only one protocol are excluded from this plot.

**Figure S2.**
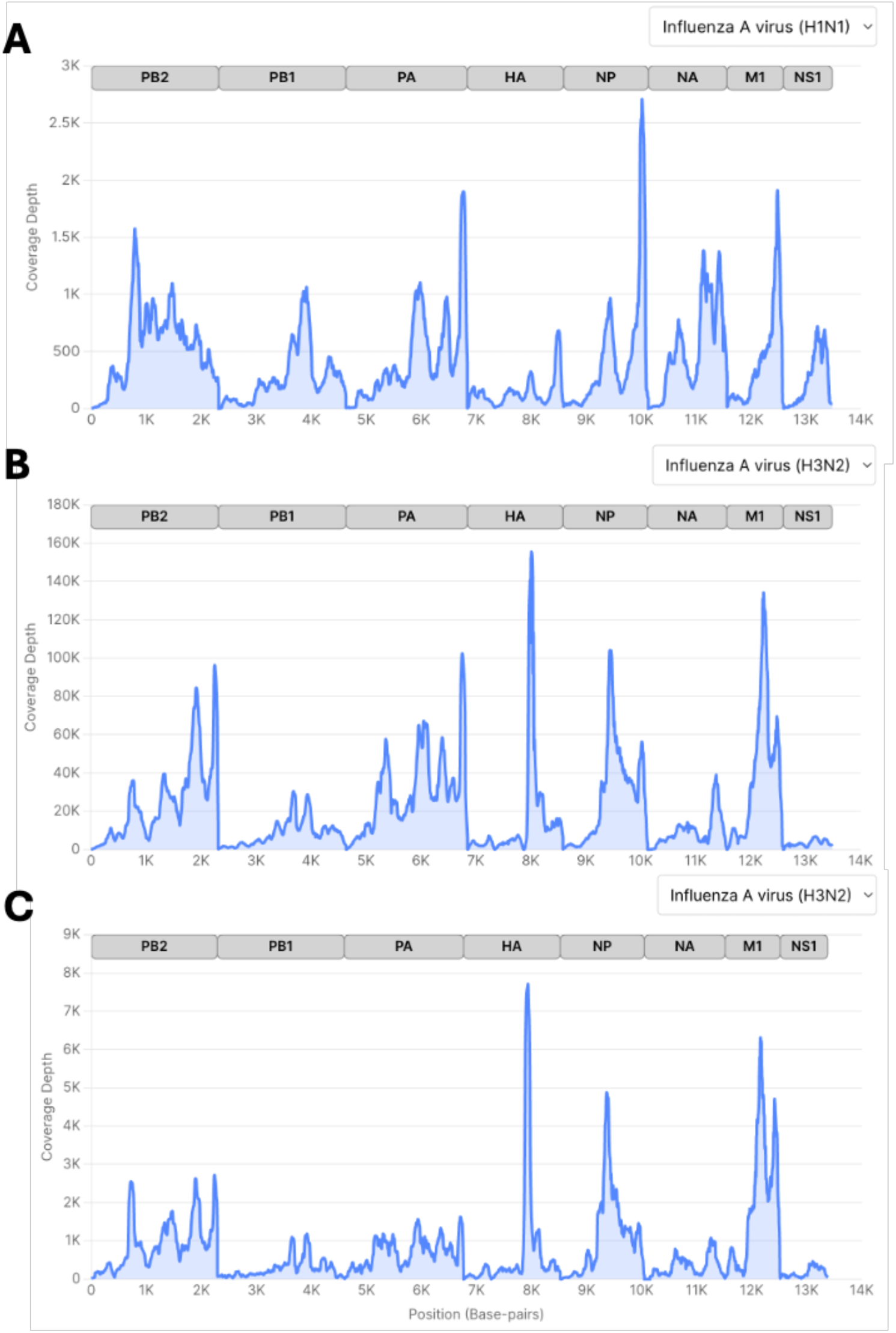
Genome-wide coverage depth across influenza gene segments in three SOC-confirmed influenza-positive clinical samples. Coverage depth (reads per position, DME+ output) is plotted against genome position (base-pairs) for the eight influenza gene segments (PB2, PB1, PA, HA, NP, NA, M1, NS1), indicated by the grey bars above each track. (A) Influenza A virus (H1N1)-positive sample. (B, C) Two independent Influenza A virus (H3N2)-positive samples. Note that the y-axis scale differs between panels (maximum depth ∼2.7K in A, ∼155K in B, ∼8K in C), reflecting sample-to-sample variation in coverage rather than a shared axis; each panel should be interpreted independently.

**Figure S3.**
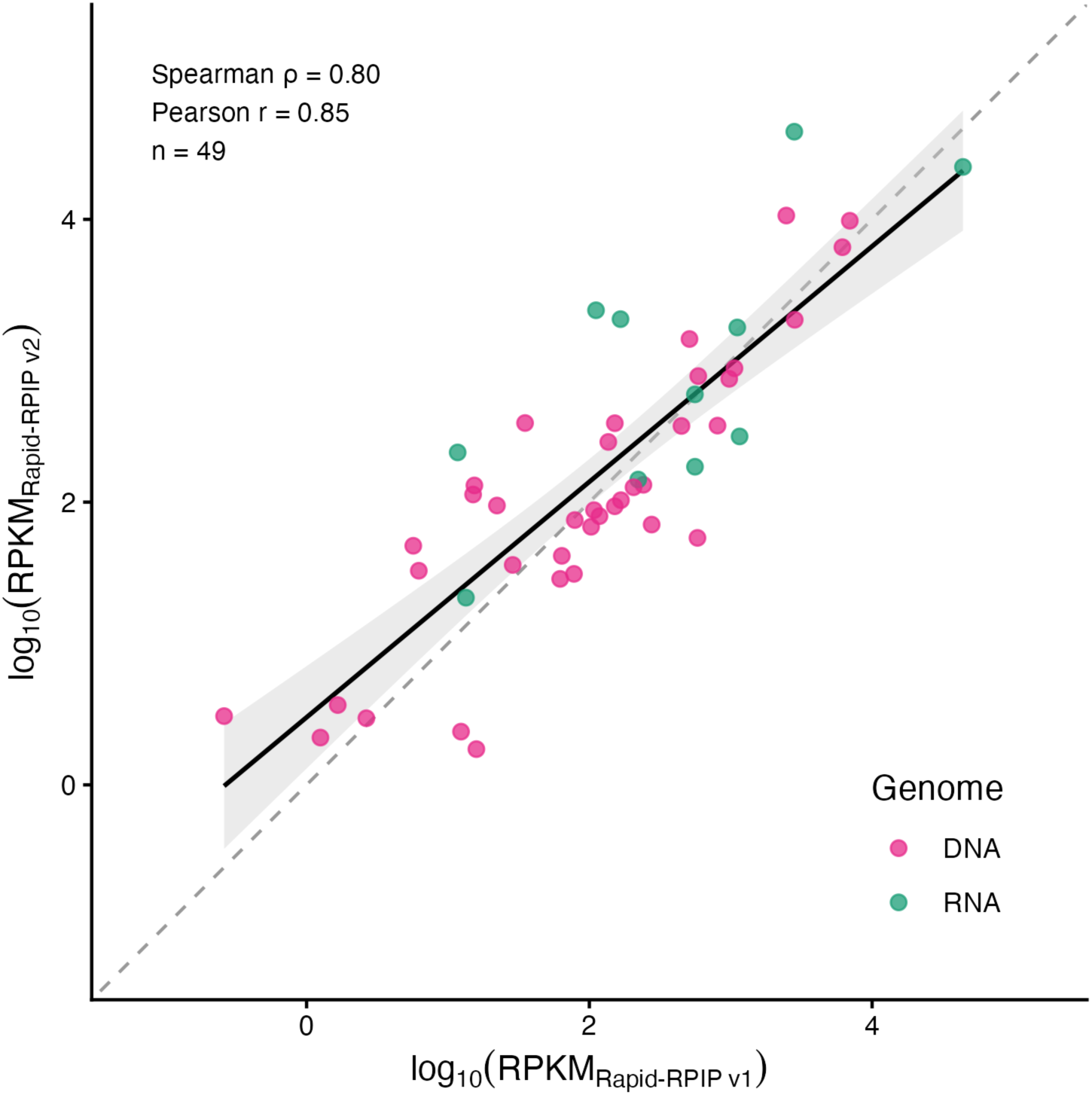
Quantitative concordance between Rapid-RPIP v1 and Rapid-RPIP v2 in clinical samples. Scatter plot of log₁₀-transformed RPKM values for organisms detected above threshold by both protocols in the same clinical sample (n=49 paired observations). Each point represents one organism–sample pair, coloured by genome type (RNA, blue; DNA, red). The dashed grey line indicates the identity line (y=x); the solid black line with grey ribbon shows the linear regression with 95% confidence interval fitted on log₁₀-RPKM values. Spearman ρ was calculated on raw RPKM; Pearson r was calculated on log₁₀-RPKM. Organisms detected by only one protocol are excluded from this plot. Note that this cohort comprises different clinical samples from those in Figure S1.

