## Supplementary material for "Same-day hybrid capture workflow for diagnosis and surveillance of respiratory pathogens": Rapid-RPIP v1 protocol updated

#### 1. Workflow

An outline of the rapid RPIP protocol is shown in Figure 1 together with the kits/reagents used at each stage.

| | |
|---|---|
| Total Nucleic Acids Extraction | Quick-DNA/RNA Pathogen Miniprep Kit |
| Rapid cDNA Synthesis | LunaScript RT SuperMix kit<br>Sequenase™ Version 2.0 DNA Polymerase |
| Sequencing Library Preparation<br>• Tagmentation<br>• Amplification<br>• Clean-up<br>• Normalisation<br><br>Target Enrichment<br>• Hybridisation<br>• Capture<br>• Amplification<br>• Clean-up | Respiratory Pathogen ID/AMR Enrichment Panel Kit |
| QC<br>• Quantification (Qubit)<br>• TapeStation | |
| Sequencing | MiniSeq Rapid Reagent kit (100) |

Figure 1. RPIP workflow outlining the different kits used.

#### 2. Reagents and kits required

##### 2.1 Reagents

| | |
|---|---|
| Human Brain Total RNA (100µg at 1 mg/mL) (Cat. AM7962) | Invitrogen |
| Human Genomic DNA: Male (100 µg at varying concentration) (Cat. G1471) | Promega |
| 2x DNA/RNA Shield (Cat. R1200-125) | Zymo Research |
| RNA from MS2 bacteriophage (Cat. 10165948001) | Merck Life Science |
| 100% Ethanol (Cat. 40-1705-25) | Severn Biotech |
| AMPure XP beads (Cat. 20119944) | Illumina |
| Molzym DNA-free water, 10x 1.7mL (05P-020-0003) | vh bio |
| NaOH 1M (Cat: S2770-100ML) | Sigma-Aldrich |
| Tris 1M pH 7 (Cat: T1819-100ML) | Sigma-Aldrich |

##### 2.2 Kits

| | |
|---|---|
| Quick-DNA/RNA Pathogen Miniprep kit (Cat. R1042) | Zymo Research |
| LunaScript RT SuperMix Kit (100 reactions) (Cat. E3010L) | New England Biolabs (NEB) |
| Sequenase™ Version 2.0 DNA Polymerase (Cat.70775Z1000UN) | ThermoFisher Scientific |
| Respiratory Pathogen ID/AMR Enrichment Panel Kit (Cat. 20047050) | Illumina |
| MiniSeq Rapid Reagent kit (100 cycles) (Cat 20044338) | Illumina |

#### 3. Labware, and equipment required

##### 3.1 Labware

| | |
|---|---|
| ZR BashingBead Lysis Tubes (0.1 & 0.5 mm) (Cat. S6012-50) | Zymo Research |
| DNA LoBind tube 1,5ml (Cat. 0030108051) | Eppendorf UK Limited |
| DNA LoBind Tube, 2mL (Cat. 0030108078) | Eppendorf UK Limited |
| 7ml Bijou Container | Any supplier |
| PCR TUBES 0,2ML 8-STRIP | Any supplier |
| Lens cleaning paper, lint free. | Any supplier |

##### 3.2 Equipment

| |
|---|
| Microbiological Safety cabinet |
| PCR Cooler 0.2 ml tubes x1 |
| Vortex x2 |
| Mini centrifuge x2 |
| Benchtop centrifuge x2 |
| FastPrep-24™ bead beating and lysis system x1 |
| Thermocycler x2 |
| DynaMag-96 Side Magnet (Cat. 10531948) x2 |
| Plate shaker x2 |
| Microheating system |

#### 4. Pre-Processing set up, cleaning and preparation of controls

##### 4.1 Pre-processing set up and cleaning

- All pre-PCR steps are to be conducted in a Microbiological Safety cabinet (MSC). The MSC, the equipment, and any plasticware (Table 1) to be used during the pre-PCR steps should be cleaned with RNAase Away, followed by 70% ethanol; the equipment and plasticware needed placed in the MSC and the MSC UV treated for 15 min prior to use.

| |
|---|
| 20µl Pipette |
| 200µl Pipette |
| 1000µl Pipette |
| 100µl Multichannel Pipette |
| 200µl Multichannel Pipette |
| Tube rack |
| DNA LoBind tube 1.5ml |
| DNA LoBind tube 2ml |
| DynaMag-96 Side Magnet |
| 7ml Bijou Container |
| PCR Tubes 0,2ml 8-Strip |

| |
|---|
| Molzym DNA-free water, (05P-020-0003) (3 aliquots per 8 samples) |
| Marker pen |
| 1 box of 20µl pipette tips |
| 2 boxes of 200µl pipette tips |
| 1 box of 1000µl pipette tips |

Table 1. Equipment required in the MSC.

- The removal of pipettes, marker pen, tube rack and other contents of the MSC should be kept to a minimum. If at any stage anything, including the sample tubes, need to be moved outside the MSC, they should be wiped down with 70% ethanol before putting them back in the MSC.
- Depending on the number of samples to be processed within a batch, follow the instructions in Table 2 for the number of negative controls (NCs) taking into consideration that a maximum of three samples/controls can be pooled for each hybridisation.

| No. of Samples | No. of Negative Controls | Total number of Samples/Controls | No. of Hybs |
|---|---|---|---|
| 1 | 2 | 3 | 1 |
| 2 | 1 | 3 | 1 |
| 3 | 3 | 6 | 2 |
| 4 | 2 | 6 | 2 |
| 5 | 1 | 6 | 2 |
| 6 | 2 | 8 | 3 |

Table 2. Experimental design for RPIP batches.

##### 4.2 Preparation of the Negative Control (NC)

1. For MS2 RNA working stock (IPC) preparation see “Appendix 1”.
2. The NC should be prepared in the MSC.
3. The reagents required for the preparation of the NC are given in Table 3

| Reagent | Instructions |
|---|---|
| Human genomic DNA | Flick the tube, centrifuge and keep on ice |
| Human brain total RNA | Thaw on ice, flick tube, briefly centrifuge and keep on ice |
| 2x DNA/RNA Shield | Vortex to mix |
| Molzym DNA-free water | Thaw at room temperature |
| MS2 RNA working stock (IPC) | Thaw on ice, invert to mix, centrifuge, and keep on ice |

Table 3. Reagents required for the preparation of the NCs

4. The NC used within this workflow consists of 500ng each of human DNA and RNA in 400 µl water mixed with 402 µl Extraction Reagent Mix.
5. Stocks of the NCs should be prepared in advance. The volumes given in points a-c below are for one NC and should be multiplied as appropriate to make multiple NCs. The NCs should be prepared in a falcon tube, mixed well and 802 µl aliquots in 1.5mL LoBind tubes stored at -80°C.
   a) Depending on the concentration indicated on the tubes, calculate the volume needed for 500ng DNA and 500ng RNA made up to 400 µl with Molzym DNA-free water (giving a concentration of 1.25ng/µl DNA/RNA).
   b) The Extraction Reagent Mix consists of 400 µl 2x DNA/RNA Shield and 2 µl of the internal positive control (IPC) MS2 RNA working stock.

On the day of processing the samples, thaw the appropriate number of aliquots at room temperature.

#### 5. DNA/RNA Extraction - Quick-DNA/RNA™ Pathogen Miniprep kit

**NOTE**

- All steps of the procedure **must be** performed in the MSC.
- When sample tubes need to be taken out of the MSC, wipe them down with ethanol before placing them back in the MSC.
- To identify potential contamination, NCs must be included in all processing batches; refer to Table 2 for the number of NCs processed in differing batch sizes. The order in which samples and controls should be arranged for different batch sizes is shown in Table 4.

| No. of samples | Tube 1 | Tube 2 | Tube 3 | Tube 4 | Tube 5 | Tube 6 | Tube 7 | Tube 8 |
|---|---|---|---|---|---|---|---|---|
| 1 | NC | S | NC | | | | | |
| 2 | S | NC | S | | | | | |
| 3 | S | NC | S | NC | S | NC | | |
| 4 | S | NC | S | S | NC | S | | |
| 5 | S | S | S or NC | S or NC | S | S | | |
| 6 | S | S | NC | S | S | S | NC | S |
| | **Hyb 1** | | | **Hyb 2** | | | **Hyb 3** | |

Table 4. Order of Samples and NCs, 'S' refers to a sample and 'NC' to a negative control. This order should be maintained when pooling samples for the hybridisation

##### 5.1 Pre-processing

1. For each sample/control, in the MSC:
   - Label 1x BashingBead Lysis Tube
   - Label and place 1x Zymo-Spin IICR column in a collection tube (both included in the Quick-Zymo extraction kit)
   - Label 1x 2mL and 1x 1.5mL LoBind tubes

   For each batch of samples, prepare a bijou with 550µL of EtOH (95-100%) per sample (this includes 10% excess EtOH). Wipe down all tubes with 70% ethanol and place them in the MSC once prepared.

| Reagent | Instructions |
|---|---|
| 2x DNA/RNA Shield | Vortex to mix |
| MS2 RNA working stock (IPC) | Thaw on ice, invert to mix, centrifuge, and keep on ice |
| DNase/RNase-Free Water | |
| Pathogen DNA/RNA Buffer | Invert to mix |
| Pathogen DNA/RNA Wash Buffer | Invert to mix |
| 95-100% EtOH | |

Table 5. Reagents used for sample pre-processing and during extraction

| Labware |
|---|
| BashingBead Lysis Tubes |
| Collection Tubes |
| Zymo-Spin IICR Columns |
| DNA LoBind Tube, 2mL |
| DNA LoBind Tube, 1.5mL |

Table 6. Labware used during extraction

2. If samples are not received in DNA/RNA Shield, prepare an adequate volume (plus 0.5x excess volume) of Extraction Reagent Mix for the number of samples (see section 4.2.5b). Add 402 µl of Extraction Reagent Mix into each empty BashingBead Lysis Tube and store them at room temperature.

3. For samples received in DNA/RNA Shield, ensure that there is a 1:1 volume of primary sample:DNA/RNA Shield. The final volume should be 800 µl sample:DNA/RNA Shield mix) plus 2 µl of the MS2 RNA working stock IPC (see 4.2).

#### 5.2 Sample Homogenisation and extraction

**NOTE**

- All steps to be performed at room temperature and centrifugation is at 10,000 rcf for 30 s, unless specified otherwise.
- See tables 4 and 5 for reagents and labware needed during extraction.

1. Retrieve all samples and controls to be tested and thaw them on ice (if frozen). Briefly vortex and centrifuge, wipe with 70% ethanol before placing on ice in the MSC.
2. Check samples and depending on sample status do one of the following:
    - For samples which are not received in DNA/RNA Shield, add 400 µl sample into the corresponding BashingBead Lysis Tube (Extraction Reagent Mix already added).
    - For samples received in DNA/RNA Shield, add 800 µl of the mixture to the corresponding BashingBead Lysis Tube and spike in with 2 µl of MS2 RNA working stock (IPC).
    - For samples received in Extraction Reagent Mix, add 802 µl of the mixture to the corresponding BashingBead Lysis Tube.
3. Remove the samples from the MSC and secure them into the sample holder adapter within the FastPrep-24 5G (or equivalent bead-beating).
4. Homogenise samples using the following settings: speed 6 m/s, two 60 s cycles with a 5 min cooling period between cycles.
5. During homogenisation, add 800 µl of Pathogen DNA/RNA Buffer to the already labelled 2.0mL tubes in the MSC.
6. Once homogenisation is complete, centrifuge the samples at 10,000 rcf for 2 min.
7. In the MSC- transfer 400 µl of the homogenate supernatant to the tube containing Pathogen DNA/RNA Buffer. Mix thoroughly by vortexing for 5 s (outside the MSC).
8. Return to the MSC. Transfer the mixture, in two steps, into a Zymo-Spin™ IICR Column in a Collection Tube and centrifuge (outside the MSC). Return to the MSC before discarding the flow-through.
9. Add **500 µl Pathogen DNA/RNA Wash Buffer** to the column and centrifuge (outside the MSC). Return to the MSC before discarding the flow-through - repeat this wash step.
10. Add 500 µl ethanol (95-100%) to the column and centrifuge (outside the MSC) for 1 min to ensure removal of any residual ethanol. Return to the MSC before discarding the collection tube and carefully transferring the column into the already labelled 1.5mL LoBind tube.
11. To elute DNA/RNA, add **50 µl DNase/RNase-Free Water** provided in the extraction kit directly to the matrix of the column, incubate for 5 min, and then centrifuge at 8,000 rcf for 1 min (outside the MSC) – place on ice in the MSC.
    - ❖ During the 5 min incubation, the cDNA synthesis reagents (refer to section 6, Table 7) can be thawed on ice in the MSC. Label one strip-tube and on ice aliquot 4 µl of LunaScript RT SuperMix for each sample.
12. Proceed **immediately** to cDNA synthesis.

### 6. cDNA synthesis and Heat Inactivation

**NOTE**

- All sample and reagent transfer steps **must be** performed in the MSC.
- All steps to be performed on ice.
- Before being placed in the MSC or if samples are taken outside of the MSC to perform some steps (e.g. centrifugation), wipe all tubes with ethanol.

| Reagent | Instructions before Use |
|---|---|
| LunaScript RT SuperMix | Place on ice. Invert to mix, centrifuge briefly, and keep on ice. |
| 5x Sequenase Buffer | Thaw on ice. Invert to mix, centrifuge briefly, and keep on ice. |
| Sequenase Dilution Buffer | Thaw on ice. Invert to mix, centrifuge briefly, and keep on ice. |
| Molzym DNA-free water | Bring to room temperature. |

| | |
|---|---|
| Sequenase enzyme | Take it out just before adding to the Master Mix. Mix by inverting and briefly centrifuge. Keep it on ice while adding to the Master Mix. |

Table 7. Storage and Preparation of reagents used for cDNA synthesis.

1. Transfer 16 µl of the extracted sample to the strip-tube containing the LunaScript Mix. Mix well by pipetting using a multichannel pipette 20 times, flicking the tubes, and briefly spinning down. Store residual extract at -80°C.
2. Place the tubes in the thermocycler and run the following program: 25°C for 2 min, 55°C for 10 min, 95°C for 1 min. While the program is running:
    - ❖ Prepare the Sequenase Master Mix (point 3 below, Table 8)
    - ❖ Place the tagmentation reagents on ice or at room temperature in the MSC (depending on the reagent – see section 7, table 9) for next step.
3. Prepare the Sequenase Master Mix and pipette well to mix. **NOTE:** add the Sequenase enzyme just before adding the Master Mix to the sample (point 4 below).

| **cDNA synthesis reagents** | **Volume (µl) of different number of Samples/Controls** | | | | | | |
|---|---|---|---|---|---|---|---|
| | **1** | **3*** | **4*** | **5*** | **6*** | **7*** | **8*** |
| 5x Sequenase Buffer | 2 | 7 | 9 | 11 | 13 | 15 | 17 |
| Molzym DNA-free water | 7.7 | 26.95 | 34.65 | 42.35 | 50.05 | 57.75 | 65.45 |
| Sequenase Dilution Buffer | 0.9 | 3.15 | 4.05 | 4.95 | 5.85 | 6.75 | 7.65 |
| Sequenase enzyme | 0.6 | 2.1 | 2.7 | 3.3 | 3.9 | 4.5 | 5.1 |
| **Total volume** (µl) | **11.2** | **39.2** | **50.4** | **61.6** | **72.8** | **84** | **95.2** |

Table 8. Reagent volumes for Sequenase Master Mix preparation. *0.5x excess volume

4. Centrifuge the 20 µl of product post 1st_LS step, return to the MSC and add 11.2 µl of the Sequenase Master Mix. Mix by flicking the tubes and briefly spin them down.
5. Place the tubes in the thermocycler and run the following program: 37°C for 8 min, 95°C for 1 min and 65°C for 9 min.
    - ❖ If proceeding to tagmentation, prepare the Tagmentation Master Mix (Tag Mix - refer to section 7.1) in the MSC during the 2nd strand/heat inactivation step and leave at room temperature.
6. Centrifuge the tubes briefly and place at room temperature in the MSC to proceed with tagmentation.

**SAFE STOPPING POINT**

Samples can be stored at -25°C to -15°C for up to 7 days.

#### 7. Illumina IDbyDNA Respiratory Pathogen ID/AMR Panel (RPIP) kit

##### NOTE

- All steps in this section use Illumina's RPIP kit.
- All steps up until PCR amplification (step 7.3) **must be** performed inside the MSC. Once the samples have been indexed and amplified, all post-PCR steps (from step 7.4) **must be** carried out on the post-PCR bench.
- Before being placed in the MSC or if samples are taken outside of the MSC to perform some steps (e.g. centrifugation, vortexing), wipe the tubes with ethanol before placing them back into the MSC.

##### 7.1 Tagment cDNA – RPIP kit

This step uses Enrichment Bead-Linked Transposomes to tagment double-stranded cDNA. The tagmentation process fragments cDNA and adds adapter sequences. After tagmentation, the fragments are purified and amplified to add index adapter sequences for dual indexing and P7 and P5 sequences for clustering.

1. Prepare the reagents shown in Table 9.

| Reagents | Instructions |
|---|---|
| EBLTL (Enrichment Bead-Linked Transposomes) | Bring to room temperature, and vortex until beads resuspended |
| EPM (Enhanced PCR Mix) | Thaw on ice. Invert to mix, centrifuge briefly |
| Indexes | Thaw at room temperature. Vortex to mix and centrifuge at 1000 x g for 1 min |
| ST2 (Stop Tagment Buffer 2) | Vortex to mix and centrifuge briefly. See point 2 below. |
| TB1 (Tagmentation Buffer 1) | Bring to room temperature. Vortex to mix |
| TWB (Tagment Wash Buffer) | Vortex to mix |

Table 9. Storage and preparation of tagmentation reagents.

2. If the ST2 buffer has precipitate:
   - Heat at 37°C for 10 min
   - Vortex until precipitate is dissolved
   - Return to room temperature
3. In a 1.5mL LoBind tube at room temperature, combine exactly the following volumes to prepare Tag Mix. Multiply each volume by the number of samples.
   - TB1 (11.5 µl)
   - EBLTL (11.5 µl)
   - Nuclease-free water (14.5 µl)

   NOTE: Volumes include reagent overage for accurate pipetting.
4. Thoroughly vortex the Tag Mix to resuspend.
5. Transfer 17.5 µl sample to a new strip-tube (multichannel if possible). Residual cDNA can be stored at -20°C.
6. Add 32.5 µl Tag Mix to each tube and pipette each sample using the multichannel pipette (set it to 40ul) and thoroughly to mix.
7. Place in a thermocycler and run the following program:
   - Pre-heated lid set to 100°C
   - Reaction volume 50 µl
   - 55°C for 5 min
   - Hold at 10°C for ~5 min

##### 7.2 Wash Tagmented cDNA

1. Briefly centrifuge the tubes.
2. Incubate at room temperature for 2 min in the MSC.
3. Add 10 µl ST2 to each tube.

4. Mix thoroughly by pipetting using the multichannel pipette 20 times – setting the pipette to 50 µl.
5. Incubate at room temperature for 5 min.
    - Prepare on ice PCR Master Mix (refer to section 7.3) and leave on ice.
6. Centrifuge briefly and place on the magnet. Wait until the liquid is clear (~3 min).
7. Remove and discard all supernatant.
8. Wash beads as follows:
    a. Remove from the magnet
    b. Add 100 µl TWB to each tube
    c. Shake on the plate shaker at 2000 rpm for 1 min
    d. Briefly centrifuge
    e. Place on the magnet and wait until the liquid is clear (~2 min)
    f. Remove and discard all supernatant with a multichannel, if possible.
9. Wash beads a **second** time.
10. Wash beads a **third** time, **skipping step f**. TWB remains in the tube to prevent over-drying.
11. Keep on the magnet and immediately proceed to Amplification step (section 7.3). Each tube contains 100 µl beads with tagmented cDNA.

##### 7.3 Amplify Tagmented cDNA

1. Combine exactly the following volumes to prepare PCR Master Mix. Multiply each volume by the number of samples.
    - 23 µl EPM
    - 23 µl Molzym DNA-free water

   NOTE: Volumes include reagent overage for accurate pipetting.
2. Thoroughly vortex the PCR Master Mix to mix.
3. Keeping the tubes on the magnet, remove and discard all TWB supernatant with a multichannel pipette.
4. With a 20 µl pipette, remove all residual TWB. Foam is normal and doesn't affect the library.
5. Remove from the magnet.
6. Add 40 µl PCR Master Mix to each tube on ice.
7. Using a new pipette tip for each well, pierce the foil covering the index adapter plate wells that you intend to use.
8. Add 10 µl UDP0XXX to each tube. Transfer volumes from the index adapter plate to the PCR tubes.
9. Cap the tubes and shake at 2000 rpm on the plate shaker for 1 min.
10. Centrifuge briefly, place in the thermocycler and run the following program.
    - Choose the preheat lid option and set to 100°C
    - Reaction volume 50 µl
    - 72°C for 3 min
    - 98°C for 3 min
    - 17 cycles of:
        - 98°C for 20 s
        - 60°C for 30 s
        - 72°C for 15 s
    - 72°C for 3 min
    - Hold at 10°C

   Total program time is ~36 min and each tube contains 50 µl beads with DNA attached.
    - On the post-PCR bench:
        - Freshly prepare 80% EtOH (at least 400 µl will be needed for each sample).
        - AMPure XP beads need to be at room temperature for 30 min; vortex and invert to mix.
        - Bring RSB to room temperature for 30 min before use; vortex to mix.

**SAFE STOPPING POINT**

Samples can be stored at -25°C to -15°C for up to 7 days. Alternatively, leave on the thermocycler for up to 24 h.

#### 7.4 Clean up Library

This step uses magnetic beads to purify the tagmented library.

**NOTE**

- **From this step onward, the workflow must be carried out in the post-PCR bench. The tubes should be removed from the thermocycler and taken to the post-PCR bench before they are opened.**
- All pipettes and the bench space to be used must be cleaned with RNase Away and 70% ethanol, prior to use.
-

| Reagents | Instructions |
|---|---|
| 100% EtOH | |
| AMPure XP beads | Let stand at room temperature at least 30 min, vortex until beads are resuspended |
| RSB (resuspension Buffer | Bring to room temperature. Vortex to mix |

Table 10. Storage and preparation of clean-up reagents.

1. Centrifuge the tubes briefly
2. Place on the magnet and wait until the liquid is clear (~2 min).
3. Transfer 45 µl supernatant (with a multichannel pipette) from each tube to a new strip-tube.
4. Vortex AMPure XP to resuspend.
5. Add 81 µl AMPure XP to each sample. Setting the multichannel pipette at 100 µl mix well by pipetting.
6. Incubate at room temperature for 5 min.
7. If needed, centrifuge the tubes briefly and place them on the magnet. Wait until the liquid is clear (~ 5 min).
   - ❖ Bring the hybridisation reagents to room temperature (refer to section 7.6, table 11) and preheat the microheating system to 50°C.

8. Remove and discard all supernatant (~ 126 µl), being careful to not take any beads.
9. Wash beads as follows.
   I. Keep on the magnet and add 175 µl fresh 80% EtOH to each sample.
   II. Wait 30 s and remove and discard all supernatant.
10. Wash beads a **second** time.
11. With a 20 µl pipette, remove all residual EtOH. Air-dry on the magnet for 2 min but do not over-dry the beads.
12. Remove from the magnet and add 17 µl RSB to each sample.
13. Close the lids and shake at 2400 rpm on plate shaker for 1 min.
14. Incubate at room temperature for 2 min.
15. Centrifuge briefly and place on the magnet; wait until the liquid is clear (~1 min).
16. Transfer 15 µl supernatant (with a multichannel pipette) from each tube to a new strip-tube.

**SAFE STOPPING POINT**

Samples can be stored at -25°C to -15°C for up to 30 days.

##### 7.5 Pre-capture preparation

1. For single-plex enrichment, transfer 7.5 µl from each sample or NC to a new strip tube.
2. For three-plex enrichment, transfer 2.5 µl from each library (sample or NC) to a new strip tube for a final volume of 7.5 µl. Mix well by pipetting and centrifuge briefly.

##### 7.6. Hybridise Probes

This step adds capture probes to samples to target regions of interest. The procedure uses enrichment reagents and oligos from the RPIP enrichment panel.

1. Prepare the reagents (Table 11).

| Reagents | Instructions |
|---|---|
| EHB2 (Enrich Hyb Buffer 2) | Bring to room temperature, and vortex to mix. See point 3 below. |
| Enrichment oligos (RPIP Panel) | Thaw at room temperature. Vortex to mix. |
| NHB2 (Hyb Buffer 2 + IDT NXT Blockers) | Thaw at room temperature. Vortex to mix. Lay in the preheated microheating system (50°C) and incubate for 5 min. See point 2 below. |

Table 11. Storage and preparation of reagents used during hybridisation

2. Mix preheated NHB2 as follows.
   I. Vortex 3 times for 10 s each time.
   II. Pipette to fully resuspend. Keep warm until use to prevent the reformation of precipitates.
3. If EHB2 or NHB2 appears crystalised or cloudy, vortex or pipette until clear.
4. Add the following volumes **in the order listed** to each hybridisation sample (7.5 µl).
   - NHB2 (12.5 µl)
   - Enrichment oligos (RPIP panel) (2.5 µl)
   - EHB2 (2.5 µl)
5. Pipette 10 times to mix, using the multichannel pipette (set to 20 µl) and centrifuge briefly.
   NOTE: EHB2 can make the reaction appear cloudy, which is normal.
6. Place in the thermocycler and run the following program:
   - Preheat lid option and set to 100°C
   - Reaction volume 25 µl
   - 95°C for 5 min
   - 18 cycles of 1 min each:
     - 94°C for the first cycle
     - Decrease 2°C per subsequent cycle
   - 58°C for 30 min

   The total program time is ~1 h.

   ❖ Bring the Capture reagents to room temperature (refer to section 7.7, table 12) and preheat the microheating system to 58°C.

##### 7.7 Capture Hybridised Probes

This step uses magnetic beads to capture probes hybridised to the targeted library fragments of interest. Heated washes remove nonspecific binding from the beads. The enriched library is then eluted from the beads.

1. Prepare the following reagents:

| Reagents | Instructions |
|---|---|
| EE1 (Enrichment Elution Buffer 1) | Thaw at room temperature and vortex to mix. |
| EEW (Enhanced Enrichment Wash) | Thaw at room temperature. Vortex to mix. |
| ET2 (Elute Target Buffer 2) | Bring to room temperature. Vortex to mix. |
| HP3 (2 N NaOH) | Thaw at room temperature. Vortex to mix. |
| SMB3 (Streptavidin Magnetic Beads 3) | Let stand for 30 min to bring to room temperature. Vortex and invert to mix. |

Table 12. Storage and preparation of reagents used to capture the hybridised probes

2. Place EEW in the preheated microheating system. Keep heated.
3. Set the thermocycler as follows:
    - If it has an incubation option, set it to 58°C.
    - If it does not have an incubation option:
      -Preheat lid option to 70°C and hold at 58°C (Reaction volume is 100 µl)

##### 7.8 Capture

1. Take the tubes from the thermocycler and centrifuge briefly.
2. Vortex SMB3 to resuspend.
3. Add 62.5 µl SMB3 to each sample.
4. Slowly pipette (with a multichannel pipette set to 70ul) until the beads are resuspended.
5. Place in the 58°C thermocycler and incubate for 15 min. Keep the program on as the thermocycler runs continuously through the capture and 4 washes.
6. Immediately after the 15 min incubation:
    a) Centrifuge the tubes briefly
    b) Place them on the magnet and wait until the liquid is clear (~2 min)
7. Remove and discard all supernatant (87.5 µl).

##### 7.9 First Wash

1. Remove from the magnet and add 50 µl preheated EEW to each sample.
2. Shake at 2400 rpm for 4 min – return unused EEW to the microheating system and keep heated.
3. Return the tubes to the 58°C thermocycler and incubate for 5 min

4. Immediately after the 5 min incubation:
    a) Centrifuge briefly
    b) Place on the magnet and wait until the liquid is clear (~1 min)
5. Remove and discard all supernatant with a multichannel pipette.

##### 7.10 Second and Third Washes

1. Remove from the magnet and add 50 µl preheated EEW to each sample.
2. Shake at 2000 rpm for 1 min – return unused EEW to the microheating system and keep heated.
3. Return the tubes to the 58°C thermocycler and incubate for 5 min.
4. Immediately after the 5 min incubation:
    a) Centrifuge briefly
    b) Place on the magnet and wait until the liquid is clear (~1 min).
5. Remove and discard all supernatant with a multichannel pipette.
6. Repeat steps 1-5 for the **third wash**.

##### 7.11 Transfer Wash

1. Remove from the magnet and add 50 µl preheated EEW to each sample.
2. Shake at 2000 rpm for 1 min.
3. Centrifuge briefly.
4. Transfer 50 µl resuspended bead solution (with a multichannel pipette) from each tube to a new strip-tube.
   The transfer minimizes residual reagents that can inhibit amplification.
5. Centrifuge briefly.
6. Return to the 58°C thermocycler and incubate for 5 min.
   - ❖ Prepare Elution Master Mix (refer to section 7.12)
   - ❖ At the same time, take out amplification reagents and thaw them on ice (refer to section 7.13, Table 13).
7. Immediately after the 5 min incubation, place on the magnet and wait until the liquid is clear (~1 min).
8. Remove and discard all supernatant with a multichannel pipette.
9. With a 20 µl pipette, remove and discard all residual EEW.
10. Immediately proceed to section 7.12 to prevent over-drying.

##### 7.12 Elution

1. Combine exactly the following volumes to prepare Elution Master Mix. Multiply each volume by the total number of samples.
   - EE1 (28.5 µl)
   - HP3 (1.5 µl)

   Volumes include reagent overage for accurate pipetting despite possible foam.
2. Vortex briefly the Elution Master Mix to mix, and then set aside at room temperature.
3. Remove the tubes from the magnet.
4. Add 23 µl Elution Master Mix to each sample.
5. Shake at 2400 rpm for 1 min.
6. Incubate at room temperature for 2 min.
7. Centrifuge briefly and place on the magnet; wait until the liquid is clear (~2 min).
8. Transfer 21 µl supernatant (with a multichannel pipette) from each sample to a new strip-tube.
9. Add 4 µl ET2 to each sample.
10. Mix thoroughly by pipetting 10 times, setting the multichannel pipette to 20 µl. Each tube contains a volume of 25 µl (Enriched sample).

**SAFE STOPPING POINT**
Samples can be stored at -25°C to -15°C for up to 7 days.

##### 7.13 Amplify Enriched Library

This step uses a 14-cycle PCR program to amplify the enriched library.

1. Prepare the following reagents (Table 13):

| Reagents | Instructions |
|---|---|
| EPM (Enhanced PCR Mix) | Thaw on ice. Invert to mix and centrifuge briefly. Keep on ice |
| PPC (PCR Primer Cocktail) | Thaw on ice. Invert to mix and centrifuge briefly. Keep on ice |

Table 13. Storage and preparation of reagents used during Amplification and Clean up of Enriched Library.

2. Centrifuge the tubes briefly.
3. Add 5 µl PPC to each sample.
4. Add 20 µl EPM to each sample.

5. Shake at 2000 rpm on plate shaker for 1 min.
6. Centrifuge briefly.
7. Place in the thermocycler and run the following program:
    - Choose the preheat lid option and set to 100°C
    - Reaction volume 50 µl
    - 98°C for 30 s
    - 14 cycles of:
        - 98°C for 10 s
        - 60°C for 30 s
        - 72°C for 30 s
    - 72°C for 5 min
    - Hold at 10°C

    Total program time is ~35 min.

    ❖ Let AMPure XP and RSB stand at room temperature for at least 30 min and freshly prepare 80% EtOH (at least 400 µl for each sample) - see section 12.14, Table 14.

    SAFE STOPPING POINT
    Samples can be stored at 2°C to 8°C for up to 2 days. Alternatively, leave on the thermocycler for up to 24 h.

##### 7.14 Clean up Enriched Library

This step uses magnetic beads to purify the enriched library.

| Reagents | Instructions |
|---|---|
| 100% EtOH | |
| AMPure XP beads | Let stand at room temperature at least 30 min, vortex until beads resuspended |
| RSB (Resuspension Buffer) | Let stand for 30 min at room temperature. Vortex and invert to mix. |

Table 14. Storage and preparation of reagents used during clean-up

1. Centrifuge the samples briefly.
2. Vortex AMPure XP to resuspend.
3. Add 90 µl AMPure XP to each sample and mix well by pipetting using a multichannel pipette, setting the pipette to 100 µl.
4. Incubate at room temperature for 5 min.
5. Centrifuge briefly.
6. Place on the magnet and wait until the liquid is clear (~4 min).
7. Remove and discard all supernatant (~140 µl) with a multichannel pipette.
8. Wash beads as follows.
    a. Keep on the magnetic stand and add 175 µl fresh 80% EtOH to each sample.
    b. Wait 30 s.
    c. Remove and discard all supernatant with a multichannel pipette.
9. Wash beads a **second** time.
10. With a 20 µl pipette, remove all residual EtOH.
11. Air-dry on the magnet for ~2 min.
12. Remove from the magnet.
13. Add 32 µl RSB to each sample.
14. Shake at 2600 rpm for 1 min.
15. Incubate at room temperature for 2 min.
16. Centrifuge briefly.
17. Place on the magnet and wait until the liquid is clear (~2 min).

18. Transfer 30 µl supernatant (with a multichannel pipette) from each tube to a new strip-tubes.

**SAFE STOPPING POINT**
Samples can be stored at -25°C to -15°C for up to 7 days.

#### 8. Setting up the Sequencing Run

##### 8.1 QC of Final Libraries

1. Check the final libraries using both of the following methods:
    - Analyse 2 µl final library with the Qubit dsDNA HS Assay kit to quantify library concentration (ng/µl).
    - Analyse 2 µl enriched library with the Agilent TapeStation HS D1000 to qualify and obtain the peak size to calculate the molarity.
    - Obtain the molarity value of the library using the following formula:

$$\frac{ng/\mu l}{660\ g/mol \times average\ library\ size} \times 10^6 = Molarity\ (nM)$$

##### 8.2 Dilute Library to the Starting and Loading Concentrations

This step dilutes libraries to the starting concentration for the MiniSeq system. After diluting to the starting concentration, libraries are ready to be denatured and diluted further to the final loading concentration.

1. Dilute the libraries to **2nM** starting concentration. Qubit (dsDNA HS Assay) to confirm the concentration.
2. Calculate the volumes required to pool and dilute each 2nM library pool to **1nM** with a volume of > 10 µL.
3. Briefly vortex and spin to collect each 1nM pool after preparation.
4. Pool 5µL of each 1nM pool into a single master pool tube to be taken forward for sequencing.
5. Qubit 2µL (dsDNA HS Assay) and tape 2 µL (Agilent TapeStation HS D1000) of the final 1nM master pool. Re-calculate the molarity of the master pool using the equation from step 8.1.
6. Use this obtained concentration to dilute and denature for sequencing.

##### 8.3 Setting up the MiniSeq

Set up the rapid MiniSeq run according to manufacturer instructions. RPIP libraries are run at a final concentration of 1.6 pM on the MiniSeq. Other Illumina sequencing kits and devices can be used to sequence the final library, but optimum loading concentration may have to be optimised.

#### Appendix 1: MS2 RNA working stock (IPC)

1. MS2 is received as an ampule of unquantified RNA in 500 µl.
2. Freeze immediately upon receipt; thaw and keep on ice at all times to minimise RNA degradation.
3. After first thaw, make a 10-fold dilution series of the neat MS2 stock to $10^{-6}$ in TE Buffer. Test this dilution series by real-time PCR (see below) to identify the dilution that gives a Ct Value between 29-31.
4. Once the appropriate dilution has been identified:
    - Make a **working** stock at the appropriate dilution and store at -20°C in 50 µl aliquots. Each aliquot is sufficient for approximately 23 samples. These aliquots are ready to use. Minimise freeze-thaw cycles (maximum five freeze-thaw cycles per aliquot). Discard any residual material thereafter.

- Make aliquots of the remaining **neat** stock and store at -20°C. These aliquots should be used to make the working stock at the appropriate dilution. The neat aliquots should only be thawed once to prepare the working stock. Discard any unused material after thawing

***MS2 Real-time PCR***

1. This information is for setting up qPCR using the QuantiNova Probe RT-PCR kit on a StepOnePlus PCR system. Protocol may need modifications when using other qPCR kits and machines.
    - Add 100ul of QN ROX Reference Dye to a new tube (1ml) of 2x QuantiNova Probe RT-PCR Master Mix. This is required for StepOnePlus PCR systems.
    - Label as 2x QuantiNova Probe RT-PCR Master Mix with "ROX" and date. Store tube at -20°C after use.
    - This only applies for QuantiNova Probe RT-PCR cat 208352.

2. Primer and probe sequences:

| **PCR target** | **Primer name** | **Sequence (5'-3')** | **Target** | **Reference** |
|---|---|---|---|---|
| MS2 bacteriophage | MS2-TM3-F | GGC TGC TCG CGG ATA CCC | 183 nt; replicase, beta subunit | (Dreier et al., 2005) |
| | MS2-TM3-R | TGA GGG AAT GTG GGA ACC G | | |
| | MS2-TM2JOE | Cy5 - ACC TCG GGT/TAO/TTC CGT CTT GCT CGT – Iowa Black RQ-Sp | | |

3. PCR master mixes:

| **MS2 (RNA)** | **Vol. for 1 reaction (µl)** |
|---|---|
| QuantiNova Probe RT-PCR Master mix with ROX dye | 13.75 |
| Forward (10 µM) | 0.75 (0.3 µM) |
| Reverse (10 µM) | 2.25 (0.9 µM) |
| Probe (10 µM) | 0.625 (0.25 µM) |
| Water | 2.375 |
| QN Probe RT-Mix | 0.25 |
| **Total Mastermix volume** | 20 |
| Volume RNA | 5 |

4. PCR cycling conditions:

| **MS2 PCR** |
|---|
| 45°C 10 minutes<br>95 °C 5 minutes<br><br>45 cycles of:<br>95 °C 5 seconds<br>60 °C 30 seconds |
