## Supplementary material for "Same-day hybrid capture workflow for diagnosis and surveillance of respiratory pathogens": Rapid-RPIP v2 protocol updated

### 1. Workflow

An outline of the rapid RPIP protocol is shown in Figure 1 together with the kits/reagents used at each stage.

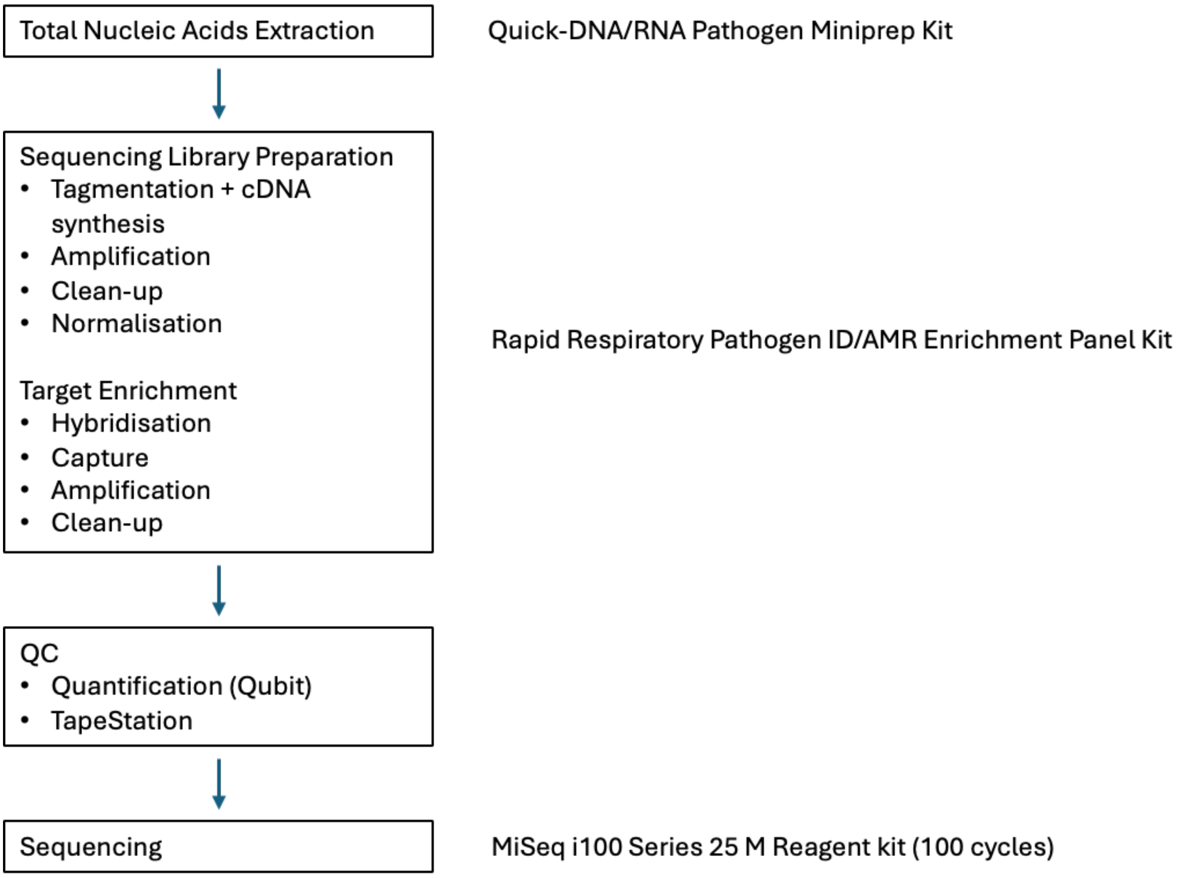


Figure 1. Rapid-RPIP v2 workflow outlining the different kits used.

### 2. Reagents and kits required

#### 2.1 Reagents

| | |
|---|---|
| Human Brain Total RNA (100µg at 1 mg/mL) (Cat. AM7962) | Invitrogen |
| Human Genomic DNA: Male (100 µg at varying concentration) (Cat. G1471) | Promega |
| 2x DNA/RNA Shield (Cat. R1200-125) | Zymo Research |
| RNA from MS2 bacteriophage (Cat. 10165948001) | Merck Life Science |
| 100% Ethanol (Cat. 40-1705-25) | Severn Biotech |
| AMPure XP beads (Cat. 20119944) | Illumina |
| Molzym DNA-free water, 10x 1.7mL (05P-020-0003) | vh bio |
| NaOH 1M (Cat: S2770-100ML) | Sigma-Aldrich |
| Tris 1M pH 7 (Cat: T1819-100ML) | Sigma-Aldrich |

#### 2.2 Kits

| | |
|---|---|
| Quick-DNA/RNA Pathogen Miniprep kit (Cat. R1042) | Zymo Research |
| Illumina Rapid Respiratory Pathogen ID/AMR RPIP Kit (Cat. 20158828) | Illumina |
| Illumina® DNA/RNA UD Indexes Set A, Tagmentation (96 Indexes, 96 Samples) (Cat. 20091654) | Illumina |
| MiSeq i100 Series 25M Reagent kit (100 cycles) (Cat. 201226567) | Illumina |

#### 6. Illumina Rapid IDbyDNA Respiratory Pathogen ID/AMR Panel kit

##### NOTE

- All steps in this section use Illumina's Rapid Respiratory pathogen ID/AMR Panel kit.
- All steps up until PCR amplification (step 6.3) **must be** performed inside the MSC. Once the samples have been indexed and amplified, all post-PCR steps (from step 6.4) **must be** carried out on the post-PCR bench.
- Before being placed in the MSC or if samples are taken outside of the MSC to perform some steps (e.g. centrifugation, vortexing), wipe the tubes with ethanol before placing them back into the MSC.

| Reagent | Instructions before Use |
|---|---|
| cDNA Buffer Mix (CBM) | Thaw at room temperature. Invert to mix, centrifuge briefly. |
| CDNA Enzyme Mix (CEM) | Thaw at room temperature. Invert to mix, centrifuge briefly. |
| Library Reducing Reagent (LRA) | Thaw at room temperature. Invert to mix, centrifuge briefly. |
| EBLTS | Bring to RT. Vortex for 30 s. |
| Stop Tagmentation Buffer (ST2) | Vortex to mix, centrifuge briefly |
| Tagmentation Wash Buffer (TWB2) | Invert to mix. |

Table 7. Preparation of reagents used for cDNA synthesis.

#### 6.1 cDNA synthesis + tagmentation

This combines cDNA synthesis and Enrichment Bead-Linked tagmentation of double-stranded cDNA. The tagmentation process simultaneously fragments cDNA and adds partial adapter sequences. After tagmentation, the fragments are purified and amplified to add index adapter sequences for dual indexing and P7 and P5 sequences for clustering.

1. Transfer 8.5 µl of the extracted sample to the strip-tube containing the pre aliquoted EPH3. Mix well by pipetting using a multichannel pipette 20 times, briefly spinning down. Residual extracted nucleic acids can be stored at -80°C.
2. Place the tubes in the thermocycler and run following program: 65°C for 5 min, hold at 4°C). While the program is running:
    - ❖ Prepare the cDNA & tagmentation master mix at room temperature (point 3 below, Table 8)
    - ❖ Once constructed, keep master mix at room temperature in the MSC.
    - ❖ Return the cold cDNA & tagmentation reagents to their relative storage locations.

3. Prepare the cDNA and tagmentation master mix and pipette well to mix. Ensure EBLTS beads are homogenised before pipetting. Beads can be briefly vortexed, or pipette mixed prior to aspiration.

| cDNA & tagmentation reagents | Volume (µl) of different number of Samples/Controls | | | | | | |
|---|---|---|---|---|---|---|---|
| | 1 | 3* | 4* | 5* | 6* | 7* | 8* |
| CBM | 29.7 | 89.1 | 118.8 | 148.5 | 178.2 | 207.9 | 237.6 |
| CEM | 2.9 | 8.7 | 11.6 | 14.5 | 17.4 | 20.3 | 23.2 |
| LRA | 3.7 | 11.1 | 14.8 | 18.5 | 22.2 | 25.9 | 29.6 |
| EBLTS | 11 | 33 | 44 | 55 | 66 | 77 | 88 |
| **Total volume** (µl) | 47.3 | 141.9 | 189.2 | 236.5 | 283.8 | 331.1 | 378.4 |

Table 8. Reagent volumes for cDNA synthesis and tagmentation. Required overage is included in these volumes

4. After EPH3 incubation, centrifuge the samples and return to the MSC before adding 43 µl of the cDNA & tagmentation master mix. Mix by pipetting at least 10 times and briefly spin down, ensuring the BLT beads are still homogenous.
5. Place the tubes in the thermocycler and run the following program: 37°C for 15 min, 55°C for 15 min hold at 4°C).
6. Centrifuge the tubes briefly and place at RT in the MSC to proceed immediately with washing of tagmented cDNA (step 6.2).

#### 6.2 Wash tagmented cDNA

1. Briefly centrifuge the tubes.
2. Incubate at room temperature for 2 min in the MSC.
3. Add 10 µl ST2 to each tube.
4. Mix thoroughly by pipetting using the multichannel pipette 20 times – setting the pipette to 50 µl.
5. Incubate at room temperature for 5 min.
    - ❖ Prepare on ice PCR Master Mix (refer to section 6.3) and leave on ice.
6. Centrifuge briefly and place on the magnet. Wait until the liquid is clear (~3min).
7. Remove and discard all supernatant.
8. Wash beads as follows:
    a. Remove from the magnet
    b. Add 100 µl TWB to each tube
    c. Shake on the plate shaker at 2000 rpm for 1min
    d. Briefly centrifuge
    e. Place on the magnet and wait until the liquid is clear (~2min)
    f. Remove and discard all supernatant with a multichannel, if possible.

9. Wash beads a **second** time, **skipping step f**. TWB remains in the tube to prevent over-drying.
10. Keep on the magnet and immediately proceed to Amplification step (section 6.3). Each tube contains 100 µl beads with tagmented cDNA.

##### 6.3 Amplify Tagmented cDNA

1. Combine exactly the following volumes to prepare PCR Master Mix. Multiply each volume by the number of samples.
    - 23 µl EPM
    - 23 µl Molzym DNA-free water

   NOTE: Volumes include reagent overage for accurate pipetting.
2. Thoroughly vortex the PCR Master Mix to mix.
3. Keeping the tubes on the magnet, remove and discard all TWB supernatant with a multichannel pipette.
4. With a 20 µl pipette, remove all residual TWB. Foam is normal and doesn't affect the library.
5. Remove from the magnet.
6. Add 40 µl PCR Master Mix to each tube on ice.
7. Using a new pipette tip for each well, pierce the foil covering the index adapter plate wells that you intend to use.
8. Add 10 µl UDP0XXX to each tube. Transfer volumes from the index adapter plate to the PCR tubes.
9. Cap the tubes and shake at 2000 rpm on the plate shaker for 1 min.
10. Centrifuge briefly, place in the thermocycler and run the following program.
    - Choose the preheat lid option and set to 100°C
    - Reaction volume 50 µl
    - 72°C for 3 min
    - 98°C for 3 min
    - 17 cycles of:
        - 98°C for 20 s
        - 60°C for 30 s
        - 72°C for 15 s
    - 72°C for 3 min
    - Hold at 10°C

   Total program time is ~36 min and each tube contains 50 µl beads with DNA attached.
    - On the post-PCR bench:
        - Freshly prepare 80% EtOH (at least 400 µl will be needed for each sample).
        - AMPure XP beads need to be at room temperature for 30 min; vortex and invert to mix.
        - Bring RSB to room temperature for 30 min before use; vortex to mix.

**SAFE STOPPING POINT**

Samples can be stored at -25°C to -15°C for up to 7 days. Alternatively, leave on the thermocycler for up to 24 h.

##### 6.4 Clean up Library

This step uses magnetic beads to purify the amplified library.

##### 6.11 Transfer Wash

1. Remove from the magnet and add 50 µl preheated EEW to each sample.
2. Shake at 2000 rpm for 1 min.
3. Centrifuge briefly.
4. Transfer 50 µl resuspended bead solution (with a multichannel pipette) from each tube to a new strip-tube.
   The transfer minimizes residual reagents that can inhibit amplification.
5. Centrifuge briefly.
6. Return to the 58°C thermocycler and incubate for 5 min.
    - ❖ Prepare Elution Master Mix (refer to section 6.12)
    - ❖ At the same time, take out amplification reagents and thaw them on ice (refer to section 6.13, Table 12).
7. Immediately after the 5 min incubation, place on the magnet and wait until the liquid is clear (~1 min).
8. Remove and discard all supernatant with a multichannel pipette.
9. With a 20 µl pipette, remove and discard all residual EEW.
10. Immediately proceed to section 6.12 to prevent over-drying.

$$\frac{ng/\mu l}{660\ g/mol \times average\ library\ size} \times 10^6 = Molarity\ (nM)$$

##### 7.2 Dilute Library to the Starting and Loading Concentrations

This step dilutes libraries to the starting concentration for the MiSeq i100 system. After diluting to the starting concentration, libraries are ready to be diluted further to the final loading concentration.

1. Dilute the libraries to **2nM** starting concentration. Qubit (dsDNA HS Assay) to confirm the concentration.
2. Calculate the volumes required to pool and dilute each 2nM library pool to **1nM** with a volume of > 10 µL.
3. Briefly vortex and spin to collect each 1nM pool after preparation.
4. Pool 5µL of each 1nM pool into a single master pool tube to be taken forward for sequencing.
5. Qubit 2µL (dsDNA HS Assay) and tape 2 µL (Agilent TapeStation HS D1000) of the final 1nM master pool. Re-calculate the molarity of the master pool using the equation from step 7.1.
6. Use this obtained concentration for sequencing.

#### 7.3 Setting up the MiSeq i100 run

Set up the Turbo MiSeq i100 run according to manufacturer instructions. RPIP libraries are run at a final concentration of 80 pM for the MiSeq i100. Other Illumina sequencing kits and devices can be used to sequence the final library.
